## Supplementary material for "Implementing Trust in Non-Small Cell Lung Cancer Diagnosis with a Conformalized Uncertainty-Aware AI Framework in Whole-Slide Images"

### ABSTRACT

Ensuring trustworthiness is fundamental to the development of artificial intelligence (AI) that is considered societally responsible, particularly in cancer diagnostics, where a misdiagnosis can have dire consequences. Current digital pathology AI models lack systematic solutions to address trustworthiness concerns arising from model limitations and data discrepancies between model deployment and development environments. To address this issue, we developed TRUECAM, a framework designed to ensure both data and model trustworthiness in non-small cell lung cancer subtyping with whole-slide images. TRUECAM integrates 1) a spectral-normalized neural Gaussian process for identifying out-of-scope inputs and 2) an ambiguity-guided elimination of tiles to filter out highly ambiguous regions, addressing data trustworthiness, as well as 3) conformal prediction to ensure controlled error rates. We systematically evaluated the framework across multiple large-scale cancer datasets, leveraging both task-specific and foundation models, illustrate that an AI model wrapped with TRUECAM significantly outperforms models that lack such guidance, in terms of classification accuracy, robustness, interpretability, and data efficiency, while also achieving improvements in fairness. These findings highlight TRUECAM as a versatile wrapper framework for digital pathology AI models with diverse architectural designs, promoting their responsible and effective applications in real-world settings.

| Dataset | | Category | Subcategories | Num. Slides | Num. Patients |
|---|---|---|---|---|---|
| TCGA (n=941) | | Race | White | 678 | 678 |
| | | | Not reported | 165 | 165 |
| | | | Black or African American | 80 | 80 |
| | | | Asian | 17 | 17 |
| | | | American Indian or Alaska Native | 1 | 1 |
| | | Sex | Male | 569 | 569 |
| | | | Female | 372 | 372 |
| CPTAC (n=416) | | Race | White | 768 | 244 |
| | | | Asian | 390 | 123 |
| | | | Other | 120 | 37 |
| | | | Black or African American | 10 | 6 |
| | | | Unknown | 9 | 3 |
| | | | Not reported | 8 | 2 |
| | | | American Indian or Alaska Native | 1 | 1 |
| | | Sex | Male | 959 | 288 |
| | | | Female | 344 | 127 |
| | | | Not reported | 3 | 1 |
| TCGA-OOD | UCS (n=57) | Race | White | 49 | 44 |
| | | | Black or African American | 9 | 9 |
| | | | Asian | 3 | 3 |
| | | | Not reported | 1 | 1 |
| | | Sex | Male | 0 | 0 |
| | | | Female | 62 | 57 |
| | UVM (n=70) | Race | White | 54 | 54 |
| | | | Not reported | 16 | 16 |
| | | Sex | Male | 41 | 41 |
| | | | Female | 29 | 29 |
| | BLCA (n=412) | Race | White | 376 | 327 |
| | | | Asian | 44 | 44 |
| | | | Black or African American | 25 | 23 |
| | | | Not reported | 24 | 18 |
| | | Sex | Male | 340 | 304 |
| | | | Female | 129 | 108 |
| | ACC (n=92) | Race | White | 80 | 78 |
| | | | Not reported | 12 | 11 |
| | | | Asian | 3 | 2 |
| | | | Black or African American | 1 | 1 |
| | | Sex | Male | 33 | 32 |
| | | | Female | 63 | 60 |

Supplementary Table 1: **An overview of datasets used in this study along with a summary of their demographic information.**

<table>
<tr><th colspan="2">Dataset</th><th>Primary site</th><th>Disease type</th></tr>
<tr><td colspan="2">TCGA<br>CPTAC</td><td>Bronchus and Lung</td><td>Adenomas and Adenocarcinomas</td></tr>
<tr><td rowspan="4">TCGA-OOD</td><td>UCS</td><td>Uterus, NOS</td><td>Complex Mixed and Stromal Neoplasms</td></tr>
<tr><td>UVM</td><td>Eye and Adnexa</td><td>Nevi and Melanomas</td></tr>
<tr><td>BLCA</td><td>Bladder</td><td>Adenomas and Adenocarcinomas<br>Epithelial Neoplasms, NOS<br>Squamous Cell Neoplasms<br>Transitional Cell Papillomas and Carcinomas</td></tr>
<tr><td>ACC</td><td>Adrenal Gland</td><td>Adenomas and Adenocarcinomas</td></tr>
</table>

Supplementary Table 2: **An overview of in-domain and out-of-domain datasets.**

| **Hyperparameter** | **Value** |
|---|---|
| Hidden layers (top) | 2 |
| Hidden layer dimension (top) | 1024 |
| Pooling (top) | Average |
| Dropout | 0.1 |
| Tile px | 299 |
| Effective magnification | 10X |
| Random horizontal flip prob | 0.5 |
| Random vertical flip prob | 0.5 |
| Random 90-degree rotation prob | 0.5 |
| Random JPEG compression (quality 50-100) prob | 0.5 |
| Random Gaussian blur (sigma 0.5-2.0) prob | 0.1 |
| Adam, $\beta$ | (0.9, 0.999) |
| Batch size | 64 |
| Max epochs | 4 |
| Weight decay | None |
| Early stopping monitor | Accuracy |
| Learning rate scheduler | ExponentialDecay |
| Initial learning rate | 0.0003 |
| Learning rate decay steps | 512 |
| Learning rate decay rate | 0.98 |
| Automatic mixed precision | tf32 |

Supplementary Table 3: **Hyperparameters used in training deep learning models built upon the Inception-v3 architecture.** Deterministic, MC Dropout, and SNGP share the same training protocols and preprocessing steps.

| **Hyperparameter** | **Value** |
|---|---|
| Tile px | 256 |
| Effective magnification | 20X |
| Batch size | 1 |
| Weight decay | 1e-2 |
| AdamW $\beta$ | (0.9, 0.999) |
| Initial learning rate | 1e-4 |
| Learning rate schedule | CosineAnnealingLR |
| Epochs | Up to 20 |

Supplementary Table 4: **Hyperparameters used in training foundation model-based ABMIL models.**

| Hyperparameter | Value |
|---|---|
| Num inducing | 1024 |
| GP scale | 1.0 |
| GP bias | 0. |
| GP kernel type | Gaussian |
| GP input normalization | False |
| GP cov discount factor | -1 |
| GP cov ridge penalty | 1.0 |
| GP output bias trainable | False |
| GP scale random features | False |
| GP random feature type | ORF |

Supplementary Table 5: **Hyperparameters used in the random Fourier feature approximated Gaussian process in SNGP for the Inception-v3 architecture**. The implementation is based on `tfm.nlp.layers.RandomFeatureGaussianProcess`.

| Hyperparameter | Value |
|---|---|
| Num inducing | 1024 |
| GP scale | 1.0 |
| GP bias | 0. |
| GP kernel type | Gaussian |
| GP input normalization | True |
| GP cov discount factor | -1 |
| GP cov ridge penalty | 1.0 |
| GP output bias trainable | False |
| GP scale random features | False |
| GP random feature type | ORF |

Supplementary Table 6: **Hyperparameters used in the random Fourier feature approximated Gaussian process in UNI-SNGP**. The implementation is based on `tfm.nlp.layers.RandomFeatureGaussianProcess`.

| Hyperparameter | Value |
|---|---|
| Num inducing | 512 (CONCH), 768 (TITAN) 1024(Prov-GigaPath) |
| GP scale | 1.0 |
| GP bias | 0. |
| GP kernel type | Linear |
| GP input normalization | False |
| GP cov discount factor | -1 |
| GP cov ridge penalty | 1.0 |
| GP output bias trainable | True |
| GP scale random features | False |
| GP random feature type | ORF |

Supplementary Table 7: **Hyperparameters used in the random Fourier feature approximated Gaussian process in CONCH-SNGP, Prov-GigaPath, TITAN**. The implementation is based on `tfm.nlp.layers.RandomFeatureGaussianProcess`.

| Model | Evaluation dataset | Tile level | | Patient level | |
|---|---|---|---|---|---|
| | | Accuracy | AUROC | Accuracy | AUROC |
| Deterministic | TCGA | 0.7312 ± 0.0069 | 0.8138 ± 0.0072 | 0.8426 ± 0.0133 | 0.9263 ± 0.0096 |
| MC Dropout | TCGA | 0.7321 ± 0.0076 | 0.8130 ± 0.0079 | 0.8423 ± 0.0154 | 0.9234 ± 0.0089 |
| SNGP | TCGA | 0.7437 ± 0.0079 | 0.8356 ± 0.0068 | 0.8748 ± 0.0098 | 0.9496 ± 0.0041 |
| SNGP-MIL | TCGA | - | - | **0.9046 ± 0.0092** | **0.9628 ± 0.0046** |
| SNGP-RE | TCGA | 0.7056 ± 0.0175 | 0.7838 ± 0.0112 | 0.7930 ± 0.0287 | 0.9099 ± 0.0138 |
| SNGP-EAT | TCGA | **0.9234 ± 0.0086** | **0.9462 ± 0.0098** | 0.9031 ± 0.0066 | 0.9608 ± 0.0053 |
| Deterministic | CPTAC | 0.5981 ± 0.0192 | 0.6563 ± 0.0282 | 0.7249 ± 0.0291 | 0.8522 ± 0.0220 |
| MC Dropout | CPTAC | 0.6134 ± 0.0188 | 0.6852 ± 0.0221 | 0.7481 ± 0.0241 | 0.8640 ± 0.0187 |
| SNGP | CPTAC | 0.6386 ± 0.0117 | 0.7214 ± 0.0158 | 0.7971 ± 0.0219 | 0.9090 ± 0.0096 |
| SNGP-MIL | CPTAC | - | - | 0.8446 ± 0.0187 | 0.9390 ± 0.0094 |
| SNGP-RE | CPTAC | 0.5556 ± 0.0260 | 0.6236 ± 0.0402 | 0.6732 ± 0.0463 | 0.8344 ± 0.0342 |
| SNGP-EAT | CPTAC | **0.8250 ± 0.0328** | **0.8949 ± 0.0182** | **0.8776 ± 0.0117** | **0.9461 ± 0.0070** |

Supplementary Table 8: **Performance of Inception-v3-based models on the internal TCGA and external CPTAC datasets**. SNGP-MIL, which only has patient-level results, is trained with the concatenation of tile-level representations to mimic ABMIL in the context of foundation models. Since SNGP-MIL follows a different training paradigm, we do not include its results in the main paper. The best-performing model for each dataset is highlighted in bold. Results are presented as the mean ± 95% confidence intervals, derived from 20 random data splits and corresponding model training sessions.

| Model | Evaluation dataset | Empirical coverage | | |
|---|---|---|---|---|
| | | $\alpha = 0.10$ | $\alpha = 0.05$ | $\alpha = 0.01$ |
| Deterministic | TCGA | 0.9000 ± 0.0000 | 0.9500 ± 0.0000 | 0.9900 ± 0.0000 |
| MC Dropout | TCGA | 0.9001 ± 0.0000 | 0.9500 ± 0.0000 | 0.9900 ± 0.0000 |
| SNGP | TCGA | 0.9000 ± 0.0000 | 0.9500 ± 0.0000 | 0.9900 ± 0.0000 |
| SNGP-RE | TCGA | 0.9000 ± 0.0000 | 0.9500 ± 0.0000 | 0.9900 ± 0.0000 |
| SNGP-EAT | TCGA | 0.9001 ± 0.0000 | 0.9501 ± 0.0000 | 0.9900 ± 0.0000 |
| Deterministic | CPTAC | 0.9000 ± 0.0000 | 0.9500 ± 0.0000 | 0.9900 ± 0.0000 |
| MC Dropout | CPTAC | 0.9000 ± 0.0000 | 0.9500 ± 0.0000 | 0.9900 ± 0.0000 |
| SNGP | CPTAC | 0.9000 ± 0.0000 | 0.9500 ± 0.0000 | 0.9900 ± 0.0000 |
| SNGP-RE | CPTAC | 0.9001 ± 0.0000 | 0.9501 ± 0.0000 | 0.9900 ± 0.0000 |
| SNGP-EAT | CPTAC | 0.9000 ± 0.0000 | 0.9500 ± 0.0000 | 0.9900 ± 0.0000 |

Supplementary Table 9: **Empirical tile-level coverage with CP applied to Inception-v3-based models across various error levels ($\alpha$).** The tile-level empirical coverage is stable and closely aligns with the desired coverage. Results are presented as the mean ± 95% confidence intervals, derived from 20 random data splits and corresponding model training sessions, with each including 500 conformal prediction evaluations based on random partitions of the calibration and testing datasets. In the following tables, unless otherwise specified, the tile-level experiments all use the same settings.

| Model | Evaluation dataset | Empirical coverage | | |
|---|---|---|---|---|
| | | $\alpha = 0.10$ | $\alpha = 0.05$ | $\alpha = 0.01$ |
| Deterministic | TCGA | 0.9061 ± 0.0008 | 0.9558 ± 0.0006 | 0.9900 ± 0.0003 |
| MC Dropout | TCGA | 0.9063 ± 0.0008 | 0.9563 ± 0.0006 | 0.9901 ± 0.0003 |
| SNGP | TCGA | 0.9060 ± 0.0008 | 0.9558 ± 0.0006 | 0.9899 ± 0.0003 |
| SNGP-MIL | TCGA | 0.9059 ± 0.0008 | 0.9550 ± 0.0006 | 0.9900 ± 0.0003 |
| SNGP-RE | TCGA | 0.9060 ± 0.0008 | 0.9560 ± 0.0006 | 0.9899 ± 0.0003 |
| SNGP-EAT | TCGA | 0.9063 ± 0.0008 | 0.9559 ± 0.0006 | 0.9902 ± 0.0003 |
| Deterministic | CPTAC | 0.9059 ± 0.0006 | 0.9551 ± 0.0005 | 0.9898 ± 0.0002 |
| MC Dropout | CPTAC | 0.9057 ± 0.0006 | 0.9548 ± 0.0005 | 0.9897 ± 0.0002 |
| SNGP | CPTAC | 0.9057 ± 0.0007 | 0.9546 ± 0.0005 | 0.9894 ± 0.0002 |
| SNGP-MIL | CPTAC | 0.9052 ± 0.0007 | 0.9536 ± 0.0005 | 0.9897 ± 0.0002 |
| SNGP-RE | CPTAC | 0.9057 ± 0.0006 | 0.9551 ± 0.0005 | 0.9898 ± 0.0002 |
| SNGP-EAT | CPTAC | 0.9057 ± 0.0007 | 0.9544 ± 0.0005 | 0.9896 ± 0.0002 |

Supplementary Table 10: **Empirical patient-level coverage with CP applied to Inception-v3-based models across various error levels ($\alpha$).** Compared to tile-level results, patient-level coverage is less stable due to the limited number of patients available for conformal prediction, with only 100 patients used for calibration in both the TCGA and CPTAC datasets. Results are presented as the mean ± 95% confidence intervals, derived from 20 random data splits and corresponding model training sessions, with each including 500 conformal prediction evaluations based on random partitions of the calibration and testing datasets.

| Model | Evaluation dataset | Average set size | | |
|---|---|---|---|---|
| | | $\alpha = 0.10$ | $\alpha = 0.05$ | $\alpha = 0.01$ |
| Deterministic | TCGA | 1.4008 ± 0.0007 | 1.6529 ± 0.0010 | 1.9246 ± 0.0003 |
| MC Dropout | TCGA | 1.3965 ± 0.0008 | 1.6432 ± 0.0011 | 1.9218 ± 0.0003 |
| SNGP | TCGA | 1.3600 ± 0.0007 | 1.5313 ± 0.0006 | 1.7731 ± 0.0006 |
| SNGP-RE | TCGA | 1.4314 ± 0.0011 | 1.6039 ± 0.0010 | 1.8332 ± 0.0007 |
| SNGP-EAT | TCGA | **0.9640 ± 0.0005** | **1.0982 ± 0.0017** | **1.6234 ± 0.0020** |
| Deterministic | CPTAC | 1.7025 ± 0.0011 | 1.8495 ± 0.0006 | 1.9695 ± 0.0001 |
| MC Dropout | CPTAC | 1.6764 ± 0.0011 | 1.8364 ± 0.0006 | 1.9668 ± 0.0001 |
| SNGP | CPTAC | 1.5711 ± 0.0010 | 1.7084 ± 0.0009 | 1.8678 ± 0.0006 |
| SNGP-RE | CPTAC | 1.6742 ± 0.0015 | 1.7849 ± 0.0013 | 1.9079 ± 0.0009 |
| SNGP-EAT | CPTAC | **1.1841 ± 0.0030** | **1.3806 ± 0.0031** | **1.7410 ± 0.0014** |

Supplementary Table 11: **Tile-level average set size with CP applied to Inception-v3-based models across various error levels ($\alpha$).** In few cases, the average set size is less than 1. This is because the DA error rate of the baseline model is lower than the desired value. In this situation, the model rejects and outputs a zero set to achieve exact coverage. When employing CP in the real-world setting, this can be turned off. The best-performing model for each setting is highlighted in bold. Results are presented as the mean ± 95% confidence intervals, derived from 20 random data splits and corresponding model training sessions, with each including 500 conformal prediction evaluations based on random partitions of the calibration and testing datasets.

| Model | Evaluation dataset | Average set size | | |
|---|---|---|---|---|
| | | $\alpha = 0.10$ | $\alpha = 0.05$ | $\alpha = 0.01$ |
| Deterministic | TCGA | 1.1456 ± 0.0017 | 1.3620 ± 0.0024 | 1.6676 ± 0.0033 |
| MC Dropout | TCGA | 1.1506 ± 0.0017 | 1.3582 ± 0.0021 | 1.6671 ± 0.0032 |
| SNGP | TCGA | 1.0698 ± 0.0013 | 1.2424 ± 0.0018 | 1.5919 ± 0.0034 |
| SNGP-MIL | TCGA | **1.0040 ± 0.0010** | 1.2398 ± 0.0034 | 1.6569 ± 0.0041 |
| SNGP-RE | TCGA | 1.1900 ± 0.0023 | 1.4156 ± 0.0031 | 1.7661 ± 0.0035 |
| SNGP-EAT | TCGA | 1.0119 ± 0.0011 | **1.1709 ± 0.0020** | **1.6364 ± 0.0042** |
| Deterministic | CPTAC | 1.4789 ± 0.0029 | 1.6843 ± 0.0023 | 1.8848 ± 0.0015 |
| MC Dropout | CPTAC | 1.4521 ± 0.0030 | 1.6702 ± 0.0025 | 1.8815 ± 0.0016 |
| SNGP | CPTAC | 1.3007 ± 0.0026 | 1.5166 ± 0.0026 | 1.7869 ± 0.0024 |
| SNGP-MIL | CPTAC | 1.2304 ± 0.0038 | 1.4768 ± 0.0037 | 1.7797 ± 0.0028 |
| SNGP-RE | CPTAC | 1.4891 ± 0.0030 | 1.6685 ± 0.0027 | 1.8603 ± 0.0020 |
| SNGP-EAT | CPTAC | **1.1312 ± 0.0021** | **1.3148 ± 0.0027** | **1.7011 ± 0.0038** |

Supplementary Table 12: **Patient-level average set size with CP applied to Inception-v3-based models across various error levels ($\alpha$).** The best-performing model for each setting is highlighted in bold. Results are presented as the mean ± 95% confidence intervals, derived from 20 random data splits and corresponding model training sessions, with each including 500 conformal prediction evaluations based on random partitions of the calibration and testing datasets.

| Model | Evaluation dataset | DA error rate | | | |
|---|---|---|---|---|---|
| | | No CP | $\alpha = 0.10$ | $\alpha = 0.05$ | $\alpha = 0.01$ |
| Deterministic | TCGA | 0.2688 ± 0.0069 | 0.1673 ± 0.0002 | 0.1468 ± 0.0004 | 0.1372 ± 0.0005 |
| MC Dropout | TCGA | 0.2679 ± 0.0076 | 0.1663 ± 0.0002 | 0.1432 ± 0.0004 | 0.1328 ± 0.0005 |
| SNGP | TCGA | 0.2563 ± 0.0079 | 0.1566 ± 0.0002 | 0.1071 ± 0.0002 | 0.0446 ± 0.0001 |
| SNGP-RE | TCGA | 0.2944 ± 0.0175 | 0.1775 ± 0.0003 | 0.1281 ± 0.0003 | 0.0628 ± 0.0003 |
| SNGP-EAT | TCGA | **0.0766 ± 0.0086** | **0.0632 ± 0.0004** | **0.0553 ± 0.0001** | **0.0287 ± 0.0002** |
| Deterministic | CPTAC | 0.4019 ± 0.0192 | 0.3488 ± 0.0014 | 0.3462 ± 0.0014 | 0.3431 ± 0.0015 |
| MC Dropout | CPTAC | 0.3866 ± 0.0188 | 0.3189 ± 0.0012 | 0.3154 ± 0.0012 | 0.3113 ± 0.0011 |
| SNGP | CPTAC | 0.3614 ± 0.0116 | 0.2369 ± 0.0006 | 0.1765 ± 0.0007 | 0.0819 ± 0.0006 |
| SNGP-RE | CPTAC | 0.4444 ± 0.0260 | 0.3259 ± 0.0017 | 0.2592 ± 0.0019 | 0.1577 ± 0.0022 |
| SNGP-EAT | CPTAC | **0.1750 ± 0.0328** | **0.1219 ± 0.0006** | **0.0858 ± 0.0004** | **0.0448 ± 0.0005** |

Supplementary Table 13: **Tile-level DA error rate before and after applying CP to Inception-v3-based models across various error levels ($\alpha$).** The best-performing model for each setting is highlighted in bold. Results are presented as the mean ± 95% confidence intervals, derived from 20 random data splits and corresponding model training sessions, with each including 500 conformal prediction evaluations based on random partitions of the calibration and testing datasets. In the "No CP" settings, only one evaluation is performed for each trained model.

| Model | Evaluation dataset | DA error rate | | | |
|---|---|---|---|---|---|
| | | No CP | $\alpha = 0.10$ | $\alpha = 0.05$ | $\alpha = 0.01$ |
| Deterministic | TCGA | 0.1574 ± 0.0133 | 0.1070 ± 0.0008 | 0.0659 ± 0.0007 | 0.0277 ± 0.0009 |
| MC Dropout | TCGA | 0.1577 ± 0.0154 | 0.1070 ± 0.0008 | 0.0647 ± 0.0007 | 0.0389 ± 0.0024 |
| SNGP | TCGA | 0.1252 ± 0.0098 | 0.0961 ± 0.0007 | 0.0558 ± 0.0007 | **0.0190 ± 0.0005** |
| SNGP-MIL | TCGA | **0.0954 ± 0.0092** | **0.0797 ± 0.0006** | 0.0559 ± 0.0006 | 0.0292 ± 0.0018 |
| SNGP-RE | TCGA | 0.2070 ± 0.0288 | 0.1135 ± 0.0009 | 0.0730 ± 0.0008 | 0.0524 ± 0.0027 |
| SNGP-EAT | TCGA | 0.0968 ± 0.0067 | 0.0810 ± 0.0006 | **0.0508 ± 0.0006** | 0.0232 ± 0.0012 |
| Deterministic | CPTAC | 0.2751 ± 0.0291 | 0.1866 ± 0.0013 | 0.1495 ± 0.0016 | 0.0913 ± 0.0026 |
| MC Dropout | CPTAC | 0.2519 ± 0.0241 | 0.1760 ± 0.0010 | 0.1399 ± 0.0011 | 0.0725 ± 0.0012 |
| SNGP | CPTAC | 0.2029 ± 0.0219 | 0.1337 ± 0.0007 | 0.0914 ± 0.0007 | 0.0518 ± 0.0021 |
| SNGP-MIL | CPTAC | 0.1554 ± 0.0187 | 0.1240 ± 0.0008 | 0.0896 ± 0.0007 | **0.0443 ± 0.0013** |
| SNGP-RE | CPTAC | 0.3268 ± 0.0463 | 0.1931 ± 0.0013 | 0.1454 ± 0.0014 | 0.1214 ± 0.0037 |
| SNGP-EAT | CPTAC | **0.1224 ± 0.0117** | **0.1039 ± 0.0006** | **0.0645 ± 0.0005** | 0.0535 ± 0.0029 |

Supplementary Table 14: **Patient-level DA error rate before and after applying CP to Inception-v3-based models across various error levels ($\alpha$).** The best-performing model for each setting is highlighted in bold. Results are presented as the mean ± 95% confidence intervals, derived from 20 random data splits and corresponding model training sessions, with each including 500 conformal prediction evaluations based on random partitions of the calibration and testing datasets. In the "No CP" settings, only one evaluation is performed for each trained model.

| Model | Uncertainty-based OOD score | | Probability-based OOD score | |
|---|---|---|---|---|
| | AUROC | AUPR | AUROC | AUPR |
| Deterministic | - | - | 0.8764 ± 0.0154 | 0.9478 ± 0.0067 |
| MC Dropout | 0.8847 ± 0.0164 | **0.9504 ± 0.0080** | 0.8835 ± 0.0168 | 0.9499 ± 0.0082 |
| SNGP | 0.8437 ± 0.0296 | 0.9080 ± 0.0130 | 0.8975 ± 0.0119 | 0.9516 ± 0.0064 |
| SNGP-EAT | **0.9161 ± 0.0353** | 0.9394 ± 0.0183 | **0.9485 ± 0.0181** | **0.9763 ± 0.0074** |

Supplementary Table 15: **Patient-level OOD detection performance of multiple Inception-v3-based models using two distinct OOD scores.** The uncertainty averaged over the top 200 tiles with the lowest uncertainty is used to calculate the slide-level uncertainty. Deterministic can only utilize the probability-based estimate to derive its OOD score. The best-performing model for each dataset is highlighted in bold. Results are presented as the mean ± 95% confidence intervals, derived from 20 random data splits and corresponding model training sessions.

| Model | $\delta$ | UCS | UVM | BLCA |
|---|---|---|---|---|
| Deterministic | 100 | 0.8440 ± 0.0376 | 0.9202 ± 0.0150 | 0.8553 ± 0.0193 |
| MC Dropout | 100 | 0.8755 ± 0.0246 | 0.9254 ± 0.0185 | 0.8668 ± 0.0196 |
| SNGP | 100 | 0.8916 ± 0.0202 | 0.9218 ± 0.0119 | 0.8911 ± 0.0131 |
| SNGP-EAT | 100 | 0.9420 ± 0.0203 | 0.9574 ± 0.0092 | 0.9324 ± 0.0382 |
| Deterministic | 200 | 0.8662 ± 0.0277 | 0.9356 ± 0.0093 | 0.8739 ± 0.0147 |
| MC Dropout | 200 | 0.8820 ± 0.0221 | 0.9362 ± 0.0154 | 0.8793 ± 0.0193 |
| SNGP | 200 | 0.8969 ± 0.0163 | 0.9319 ± 0.0095 | 0.8917 ± 0.0152 |
| SNGP-EAT | 200 | 0.9630 ± 0.0127 | 0.9537 ± 0.0104 | 0.9493 ± 0.0271 |
| Deterministic | 300 | 0.8723 ± 0.0245 | 0.9459 ± 0.0074 | 0.8848 ± 0.0118 |
| MC Dropout | 300 | 0.8828 ± 0.0212 | 0.9440 ± 0.0135 | 0.8881 ± 0.0176 |
| SNGP | 300 | 0.8993 ± 0.0155 | 0.9379 ± 0.0085 | 0.8935 ± 0.0189 |
| SNGP-EAT | 300 | 0.9646 ± 0.0124 | 0.9440 ± 0.0126 | 0.9508 ± 0.0199 |

| Model | $\delta$ | ACC | Overall |
|---|---|---|---|
| Deterministic | 100 | 0.7864 ± 0.0509 | 0.8514 ± 0.0214 |
| MC Dropout | 100 | 0.8254 ± 0.0352 | 0.8681 ± 0.0188 |
| SNGP | 100 | 0.8750 ± 0.0276 | 0.8922 ± 0.0129 |
| SNGP-EAT | 100 | 0.9270 ± 0.0237 | **0.9353 ± 0.0267** |
| Deterministic | 200 | 0.8488 ± 0.0320 | 0.8764 ± 0.0154 |
| MC Dropout | 200 | 0.8629 ± 0.0250 | 0.8835 ± 0.0168 |
| SNGP | 200 | 0.8977 ± 0.0190 | 0.8975 ± 0.0119 |
| SNGP-EAT | 200 | 0.9315 ± 0.0188 | **0.9485 ± 0.0181** |
| Deterministic | 300 | 0.8826 ± 0.0205 | 0.8902 ± 0.0120 |
| MC Dropout | 300 | 0.8878 ± 0.0179 | 0.8938 ± 0.0149 |
| SNGP | 300 | 0.9013 ± 0.0162 | 0.9001 ± 0.0135 |
| SNGP-EAT | 300 | 0.9199 ± 0.0211 | **0.9468 ± 0.0137** |

Supplementary Table 16: **Assessment of the impact of using the top $\delta$ tiles to calculate probability-based OOD scores on OOD detection performance, measured by AUROC.** The TCGA calibration and testing datasets were combined as the In-D data, while TCGA-OOD was used as the OOD data. Performance was evaluated under two conditions: 1) considering OOD slides from a single non-NSCLC cancer type, and 2) including slides from all cancer types (represented in the "Overall" column). The best-performing model for each setting is highlighted in bold. SNGP-EAT with $\delta$=200 achieved the highest performance in OOD detection.

| Model | Evaluation dataset | Without DSC control | | With DSC control | |
|---|---|---|---|---|---|
| | | Accuracy | AUROC | Accuracy | AUROC |
| Deterministic | CPTAC | 0.5981 ± 0.0192 | 0.6563 ± 0.0282 | 0.6188 ± 0.0235 | 0.6744 ± 0.0311 |
| MC Dropout | CPTAC | 0.6134 ± 0.0188 | 0.6852 ± 0.0221 | 0.6363 ± 0.0239 | 0.7078 ± 0.0238 |
| SNGP | CPTAC | 0.6386 ± 0.0117 | 0.7214 ± 0.0158 | 0.6643 ± 0.0150 | 0.7490 ± 0.0176 |
| SNGP-EAT | CPTAC | 0.8250 ± 0.0328 | 0.8949 ± 0.0182 | **0.8873 ± 0.0221** | **0.9198 ± 0.0137** |

Supplementary Table 17: **Impact assessment of OOD detection-based distribution shift control (DSC) on tile-level classification performance in terms of accuracy and AUROC with respect to the external CPTAC dataset when the FPR for OOD detection was set at 0.2.** The results in this table correspond to Fig. 5e. Distribution shift control achieved a significant boost in the NSCLC subtyping accuracy and AUROC when applying models trained on TCGA to CPTAC, and bridged the performance gap due to distribution shift. See Supplementary Table 8 for model performance evaluated on TCGA. The best-performing model for each setting is highlighted in bold, and results are presented with mean ± 95% CI from 20 experimental replicates.

| Model | Evaluation dataset | Without DSC control | | With DSC control | |
|---|---|---|---|---|---|
| | | Accuracy | AUROC | Accuracy | AUROC |
| Deterministic | CPTAC | 0.7249 ± 0.0291 | 0.8522 ± 0.0220 | 0.7703 ± 0.0270 | 0.8303 ± 0.0278 |
| MC Dropout | CPTAC | 0.7481 ± 0.0241 | 0.8640 ± 0.0187 | 0.7878 ± 0.0307 | 0.8441 ± 0.0263 |
| SNGP | CPTAC | 0.7971 ± 0.0219 | 0.9090 ± 0.0096 | 0.8411 ± 0.0199 | 0.9095 ± 0.0137 |
| SNGP-EAT | CPTAC | 0.8776 ± 0.0117 | 0.9461 ± 0.0070 | **0.9337 ± 0.0109** | **0.9572 ± 0.0081** |

Supplementary Table 18: **Impact assessment of OOD detection-based distribution shift control (DSC) on patient-level classification performance in terms of accuracy and AUROC with respect to the external CPTAC dataset when the FPR for OOD detection was set at 0.2.** The results in this table correspond to Fig. 5f. Distribution shift control achieved a significant boost in the NSCLC subtyping accuracy and AUROC when applying models trained on TCGA to CPTAC, and bridged the performance gap due to distribution shift. See Supplementary Table 8 for model performance evaluated on TCGA. The best-performing model for each setting is highlighted in bold, and results are presented with mean ± 95% CI from 20 experimental replicates.

| Model | Evaluation dataset | $\alpha$ | Without DSC control | With DSC control |
|---|---|---|---|---|
| | | | Average set size | Average set size |
| Deterministic | CPTAC | 0.10 | 1.7025 ± 0.0011 | 1.7011 ± 0.0011 |
| MC Dropout | CPTAC | 0.10 | 1.6764 ± 0.0011 | 1.6747 ± 0.0011 |
| SNGP | CPTAC | 0.10 | 1.5711 ± 0.0010 | 1.5378 ± 0.0012 |
| SNGP-EAT | CPTAC | 0.10 | 1.1841 ± 0.0030 | **1.0676 ± 0.0022** |
| Deterministic | CPTAC | 0.05 | 1.8495 ± 0.0006 | 1.8493 ± 0.0006 |
| MC Dropout | CPTAC | 0.05 | 1.8364 ± 0.0006 | 1.8357 ± 0.0006 |
| SNGP | CPTAC | 0.05 | 1.7084 ± 0.0009 | 1.6825 ± 0.0011 |
| SNGP-EAT | CPTAC | 0.05 | 1.3806 ± 0.0031 | **1.2670 ± 0.0031** |
| Deterministic | CPTAC | 0.01 | 1.9695 ± 0.0001 | 1.9696 ± 0.0001 |
| MC Dropout | CPTAC | 0.01 | 1.9668 ± 0.0001 | 1.9668 ± 0.0001 |
| SNGP | CPTAC | 0.01 | 1.8678 ± 0.0006 | 1.8549 ± 0.0007 |
| SNGP-EAT | CPTAC | 0.01 | 1.7410 ± 0.0014 | **1.7016 ± 0.0018** |

Supplementary Table 19: **Impact assessment of OOD detection-based distribution shift control (DSC) on tile-level CP performance in terms of average set size with respect to the external CPTAC dataset when the FPR for OOD detection was set at 0.2.** The results in this table correspond to Fig. 5i. Distribution shift control achieved a significant boost in the NSCLC subtyping average set size when applying models trained on TCGA to CPTAC, and bridged the performance gap due to distribution shift. See Supplementary Table 8 for model performance evaluated on TCGA. The best-performing model for each setting is highlighted in bold, and results are presented with mean ± 95% CI from 20 experimental replicates, with each including 500 conformal prediction evaluations based on random partitions of the calibration and testing datasets.

| Model | Evaluation dataset | $\alpha$ | Without DSC control | With DSC control |
|---|---|---|---|---|
| | | | Average set size | Average set size |
| Deterministic | CPTAC | 0.10 | 1.4789 ± 0.0029 | 1.4377 ± 0.0032 |
| MC Dropout | CPTAC | 0.10 | 1.4521 ± 0.0030 | 1.4062 ± 0.0033 |
| SNGP | CPTAC | 0.10 | 1.3007 ± 0.0026 | 1.2307 ± 0.0029 |
| SNGP-EAT | CPTAC | 0.10 | 1.1312 ± 0.0021 | **0.9812 ± 0.0015** |
| Deterministic | CPTAC | 0.05 | 1.6843 ± 0.0023 | 1.6590 ± 0.0025 |
| MC Dropout | CPTAC | 0.05 | 1.6702 ± 0.0025 | 1.6443 ± 0.0027 |
| SNGP | CPTAC | 0.05 | 1.5166 ± 0.0026 | 1.4741 ± 0.0029 |
| SNGP-EAT | CPTAC | 0.05 | 1.3148 ± 0.0027 | **1.1702 ± 0.0033** |
| Deterministic | CPTAC | 0.01 | 1.8848 ± 0.0015 | 1.8748 ± 0.0016 |
| MC Dropout | CPTAC | 0.01 | 1.8815 ± 0.0016 | 1.8713 ± 0.0017 |
| SNGP | CPTAC | 0.01 | 1.7869 ± 0.0024 | 1.7728 ± 0.0026 |
| SNGP-EAT | CPTAC | 0.01 | 1.7011 ± 0.0038 | **1.6638 ± 0.0047** |

Supplementary Table 20: **Impact assessment of OOD detection-based distribution shift control (DSC) on patient-level CP performance in terms of average set size with respect to the external CPTAC dataset when the FPR for OOD detection was set at 0.2.** The results in this table correspond to Fig. 5j. Distribution shift control achieved a significant boost in the NSCLC subtyping average set size when applying models trained on TCGA to CPTAC, and bridged the performance gap due to distribution shift. See Supplementary Table 8 for model performance evaluated on TCGA. The best-performing model for each setting is highlighted in bold, and results are presented with mean ± 95% CI from 20 experimental replicates, with each including 500 conformal prediction evaluations based on random partitions of the calibration and testing datasets.

| Model | Metric | Female | Male | Fairness gap |
|---|---|---|---|---|
| Deterministic | Accuracy | 0.8347 ± 0.0228 | 0.8472 ± 0.0158 | 0.0454 ± 0.0183 |
| MC Dropout | Accuracy | 0.8268 ± 0.0240 | 0.8517 ± 0.0184 | 0.0496 ± 0.0203 |
| SNGP | Accuracy | 0.8753 ± 0.0148 | 0.8746 ± 0.0125 | **0.0312 ± 0.0117** |
| SNGP-MIL | Accuracy | 0.9323 ± 0.0108 | 0.8888 ± 0.0110 | 0.0441 ± 0.0118 |
| SNGP-RE | Accuracy | 0.7629 ± 0.0474 | 0.8114 ± 0.0325 | 0.1045 ± 0.0341 |
| SNGP-EAT | Accuracy | 0.9278 ± 0.0127 | 0.8895 ± 0.0065 | 0.0421 ± 0.0104 |
| Deterministic | AUROC | 0.9240 ± 0.0171 | 0.9168 ± 0.0110 | 0.0286 ± 0.0099 |
| MC Dropout | AUROC | 0.9209 ± 0.0148 | 0.9144 ± 0.0119 | 0.0358 ± 0.0089 |
| SNGP | AUROC | 0.9535 ± 0.0088 | 0.9400 ± 0.0073 | **0.0231 ± 0.0081** |
| SNGP-MIL | AUROC | 0.9767 ± 0.0067 | 0.9496 ± 0.0068 | 0.0282 ± 0.0083 |
| SNGP-RE | AUROC | 0.9199 ± 0.0200 | 0.8959 ± 0.0154 | 0.0388 ± 0.0141 |
| SNGP-EAT | AUROC | 0.9731 ± 0.0064 | 0.9483 ± 0.0087 | 0.0270 ± 0.0090 |

Supplementary Table 21: **Sexual fairness gaps of Inception-v3-based models in classification performance on the TCGA testing set.** The best-performing model for each dataset is highlighted in bold. Results are presented as the mean ± 95% confidence intervals, derived from 20 random data splits and corresponding model training sessions.

| Model | Metric | Female | Male | Fairness gap |
|---|---|---|---|---|
| Deterministic | Accuracy | 0.7413 ± 0.0417 | 0.7167 ± 0.0248 | 0.0471 ± 0.0120 |
| MC Dropout | Accuracy | 0.7693 ± 0.0381 | 0.7378 ± 0.0198 | 0.0545 ± 0.0136 |
| SNGP | Accuracy | 0.8232 ± 0.0289 | 0.7849 ± 0.0200 | 0.0456 ± 0.0101 |
| SNGP-MIL | Accuracy | 0.8756 ± 0.0219 | 0.8307 ± 0.0181 | **0.0450 ± 0.0099** |
| SNGP-RE | Accuracy | 0.6614 ± 0.0732 | 0.6776 ± 0.0363 | 0.0848 ± 0.0184 |
| SNGP-EAT | Accuracy | 0.9122 ± 0.0139 | 0.8627 ± 0.0118 | 0.0495 ± 0.0086 |
| Deterministic | AUROC | 0.8865 ± 0.0180 | 0.8330 ± 0.0247 | 0.0536 ± 0.0094 |
| MC Dropout | AUROC | 0.8954 ± 0.0158 | 0.8459 ± 0.0219 | 0.0495 ± 0.0126 |
| SNGP | AUROC | 0.9371 ± 0.0095 | 0.8941 ± 0.0108 | 0.0430 ± 0.0068 |
| SNGP-MIL | AUROC | 0.9663 ± 0.0073 | 0.9250 ± 0.0110 | 0.0413 ± 0.0057 |
| SNGP-RE | AUROC | 0.8699 ± 0.0306 | 0.8137 ± 0.0376 | 0.0562 ± 0.0138 |
| SNGP-EAT | AUROC | 0.9693 ± 0.0041 | 0.9358 ± 0.0088 | **0.0336 ± 0.0074** |

Supplementary Table 22: **Sexual fairness gaps of Inception-v3-based models in classification performance on the CPTAC testing set.** The best-performing model for each dataset is highlighted in bold. Results are presented as the mean ± 95% confidence intervals, derived from 20 random data splits and corresponding model training sessions.

| Model | Metric | Not reported | Others | White | Fairness gap |
|---|---|---|---|---|---|
| Deterministic | Accuracy | 0.9057 ± 0.0266 | 0.7906 ± 0.0433 | 0.8350 ± 0.0129 | 0.1487 ± 0.0338 |
| MC Dropout | Accuracy | 0.9012 ± 0.0210 | 0.7879 ± 0.0329 | 0.8345 ± 0.0167 | 0.1253 ± 0.0254 |
| SNGP | Accuracy | 0.9302 ± 0.0192 | 0.8117 ± 0.0315 | 0.8691 ± 0.0117 | 0.1284 ± 0.0372 |
| SNGP-MIL | Accuracy | 0.9295 ± 0.0182 | 0.8434 ± 0.0285 | 0.9069 ± 0.0119 | 0.1013 ± 0.0323 |
| SNGP-RE | Accuracy | 0.8738 ± 0.0294 | 0.7602 ± 0.0411 | 0.7776 ± 0.0372 | 0.1627 ± 0.0400 |
| SNGP-EAT | Accuracy | 0.9340 ± 0.0183 | 0.8505 ± 0.0203 | 0.9025 ± 0.0075 | **0.0921 ± 0.0235** |
| Deterministic | AUROC | 0.9414 ± 0.0240 | 0.8805 ± 0.0312 | 0.9271 ± 0.0098 | 0.1018 ± 0.0235 |
| MC Dropout | AUROC | 0.9343 ± 0.0269 | 0.8863 ± 0.0293 | 0.9232 ± 0.0096 | 0.0941 ± 0.0263 |
| SNGP | AUROC | 0.9557 ± 0.0186 | 0.9129 ± 0.0230 | 0.9522 ± 0.0054 | **0.0753 ± 0.0235** |
| SNGP-MIL | AUROC | 0.9608 ± 0.0154 | 0.9095 ± 0.0364 | 0.9701 ± 0.0033 | 0.0781 ± 0.0343 |
| SNGP-RE | AUROC | 0.9165 ± 0.0347 | 0.8765 ± 0.0263 | 0.9160 ± 0.0140 | 0.1023 ± 0.0220 |
| SNGP-EAT | AUROC | 0.9752 ± 0.0092 | 0.8987 ± 0.0411 | 0.9672 ± 0.0066 | 0.0859 ± 0.0408 |

Supplementary Table 23: **Racial fairness gaps of Inception-v3-based models in classification performance on the TCGA testing set.** The best-performing model for each dataset is highlighted in bold. Results are presented as the mean ± 95% confidence intervals, derived from 20 random data splits and corresponding model training sessions.

| Model | Metric | Asian | Others | White | Fairness gap |
|---|---|---|---|---|---|
| Deterministic | Accuracy | 0.5756 ± 0.0572 | 0.7224 ± 0.0359 | 0.8006 ± 0.0182 | 0.2282 ± 0.0444 |
| MC Dropout | Accuracy | 0.6012 ± 0.0571 | 0.7531 ± 0.0358 | 0.8211 ± 0.0132 | 0.2272 ± 0.0494 |
| SNGP | Accuracy | 0.7126 ± 0.0386 | 0.8061 ± 0.0331 | 0.8379 ± 0.0150 | 0.1348 ± 0.0260 |
| SNGP-MIL | Accuracy | 0.8272 ± 0.0472 | 0.8408 ± 0.0221 | 0.8541 ± 0.0117 | 0.0894 ± 0.0271 |
| SNGP-RE | Accuracy | 0.5276 ± 0.0938 | 0.6520 ± 0.0659 | 0.7508 ± 0.0363 | 0.2763 ± 0.0551 |
| SNGP-EAT | Accuracy | 0.8707 ± 0.0218 | 0.8673 ± 0.0167 | 0.8832 ± 0.0107 | **0.0496 ± 0.0121** |
| Deterministic | AUROC | 0.9286 ± 0.0162 | 0.8793 ± 0.0213 | 0.8568 ± 0.0133 | 0.0795 ± 0.0138 |
| MC Dropout | AUROC | 0.9311 ± 0.0139 | 0.9003 ± 0.0192 | 0.8652 ± 0.0110 | 0.0733 ± 0.0111 |
| SNGP | AUROC | 0.9468 ± 0.0097 | 0.9398 ± 0.0139 | 0.8950 ± 0.0113 | 0.0622 ± 0.0099 |
| SNGP-MIL | AUROC | 0.9516 ± 0.0128 | 0.9541 ± 0.0092 | 0.9256 ± 0.0100 | 0.0397 ± 0.0089 |
| SNGP-RE | AUROC | 0.9253 ± 0.0185 | 0.8546 ± 0.0407 | 0.8519 ± 0.0279 | 0.0956 ± 0.0247 |
| SNGP-EAT | AUROC | 0.9629 ± 0.0074 | 0.9492 ± 0.0103 | 0.9321 ± 0.0089 | **0.0360 ± 0.0059** |

Supplementary Table 24: **Racial fairness gaps of Inception-v3-based models in classification performance on the CPTAC testing set.** The best-performing model for each dataset is highlighted in bold. Results are presented as the mean ± 95% confidence intervals, derived from 20 random data splits and corresponding model training sessions.

| Model | $\alpha$ | **Average set size across racial groups** | | | |
|---|---|---|---|---|---|
| | | Not reported | Others | White | Fairness gap |
| Deterministic | 0.10 | 1.0871 ± 0.0017 | 1.1790 ± 0.0030 | 1.1565 ± 0.0018 | 0.1483 ± 0.0019 |
| MC Dropout | 0.10 | 1.0800 ± 0.0015 | 1.1795 ± 0.0029 | 1.1660 ± 0.0019 | 0.1582 ± 0.0020 |
| SNGP | 0.10 | 1.0252 ± 0.0009 | 1.0970 ± 0.0026 | 1.0775 ± 0.0015 | 0.1190 ± 0.0018 |
| SNGP-MIL | 0.10 | 1.0017 ± 0.0005 | 1.0130 ± 0.0021 | 1.0032 ± 0.0012 | **0.0755 ± 0.0015** |
| SNGP-RE | 0.10 | 1.1059 ± 0.0021 | 1.1964 ± 0.0032 | 1.2116 ± 0.0026 | 0.1741 ± 0.0022 |
| SNGP-EAT | 0.10 | 1.0048 ± 0.0009 | 1.0187 ± 0.0022 | 1.0132 ± 0.0012 | 0.0857 ± 0.0015 |
| Deterministic | 0.05 | 1.2273 ± 0.0026 | 1.4198 ± 0.0040 | 1.3890 ± 0.0026 | 0.2636 ± 0.0027 |
| MC Dropout | 0.05 | 1.2135 ± 0.0025 | 1.4152 ± 0.0034 | 1.3886 ± 0.0023 | 0.2716 ± 0.0027 |
| SNGP | 0.05 | 1.1151 ± 0.0017 | 1.3280 ± 0.0033 | 1.2631 ± 0.0020 | 0.2528 ± 0.0026 |
| SNGP-MIL | 0.05 | 1.1304 ± 0.0029 | 1.2967 ± 0.0046 | 1.2604 ± 0.0036 | 0.2145 ± 0.0028 |
| SNGP-RE | 0.05 | 1.2471 ± 0.0033 | 1.4367 ± 0.0039 | 1.4573 ± 0.0033 | 0.2809 ± 0.0025 |
| SNGP-EAT | 0.05 | 1.0879 ± 0.0019 | 1.2307 ± 0.0032 | 1.1842 ± 0.0021 | **0.1896 ± 0.0022** |
| Deterministic | 0.01 | 1.4889 ± 0.0042 | 1.7339 ± 0.0041 | 1.7060 ± 0.0033 | 0.2950 ± 0.0027 |
| MC Dropout | 0.01 | 1.4916 ± 0.0041 | 1.7193 ± 0.0039 | 1.7074 ± 0.0031 | 0.2870 ± 0.0027 |
| SNGP | 0.01 | 1.3776 ± 0.0040 | 1.6422 ± 0.0039 | 1.6418 ± 0.0035 | 0.3339 ± 0.0027 |
| SNGP-MIL | 0.01 | 1.5312 ± 0.0058 | 1.7043 ± 0.0044 | 1.6826 ± 0.0038 | 0.2484 ± 0.0032 |
| SNGP-RE | 0.01 | 1.6244 ± 0.0047 | 1.8083 ± 0.0039 | 1.7972 ± 0.0034 | 0.2353 ± 0.0028 |
| SNGP-EAT | 0.01 | 1.5327 ± 0.0051 | 1.6675 ± 0.0049 | 1.6600 ± 0.0042 | **0.2282 ± 0.0026** |

Supplementary Table 25: **Racial fairness gaps of Inception-v3-based models in average set size on the TCGA testing set with respect to three different error levels ($\alpha$).** The best-performing model for each setting is highlighted in bold. Results are presented as the mean ± 95% confidence intervals, derived from 20 random data splits and corresponding model training sessions, with each including 500 conformal prediction evaluations based on random partitions of the calibration and testing datasets.

| Model | $\alpha$ | **Average set size across racial groups** | | | |
|---|---|---|---|---|---|
| | | Asian | Others | Whites | Fairness gap |
| Deterministic | 0.10 | 1.5424 ± 0.0026 | 1.5343 ± 0.0032 | 1.4357 ± 0.0032 | 0.1495 ± 0.0011 |
| MC Dropout | 0.10 | 1.5190 ± 0.0028 | 1.5084 ± 0.0034 | 1.4071 ± 0.0032 | 0.1509 ± 0.0013 |
| SNGP | 0.10 | 1.3659 ± 0.0031 | 1.3590 ± 0.0034 | 1.2561 ± 0.0023 | 0.1361 ± 0.0013 |
| SNGP-MIL | 0.10 | 1.3323 ± 0.0055 | 1.2392 ± 0.0042 | 1.1773 ± 0.0030 | 0.1691 ± 0.0025 |
| SNGP-RE | 0.10 | 1.5155 ± 0.0029 | 1.5880 ± 0.0032 | 1.4559 ± 0.0033 | 0.1613 ± 0.0015 |
| SNGP-EAT | 0.10 | 1.1891 ± 0.0029 | 1.1444 ± 0.0026 | 1.0993 ± 0.0016 | **0.1086 ± 0.0013** |
| Deterministic | 0.05 | 1.7269 ± 0.0018 | 1.7492 ± 0.0026 | 1.6498 ± 0.0027 | **0.1320 ± 0.0011** |
| MC Dropout | 0.05 | 1.7236 ± 0.0019 | 1.7401 ± 0.0027 | 1.6293 ± 0.0029 | 0.1465 ± 0.0012 |
| SNGP | 0.05 | 1.5813 ± 0.0026 | 1.6113 ± 0.0032 | 1.4650 ± 0.0026 | 0.1664 ± 0.0012 |
| SNGP-MIL | 0.05 | 1.6375 ± 0.0050 | 1.5224 ± 0.0039 | 1.3867 ± 0.0031 | 0.2616 ± 0.0024 |
| SNGP-RE | 0.05 | 1.6834 ± 0.0025 | 1.7766 ± 0.0025 | 1.6393 ± 0.0031 | 0.1649 ± 0.0015 |
| SNGP-EAT | 0.05 | 1.3958 ± 0.0034 | 1.3739 ± 0.0034 | 1.2621 ± 0.0025 | 0.1638 ± 0.0015 |
| Deterministic | 0.01 | 1.8942 ± 0.0013 | 1.9482 ± 0.0014 | 1.8673 ± 0.0017 | **0.0934 ± 0.0009** |
| MC Dropout | 0.01 | 1.8890 ± 0.0012 | 1.9452 ± 0.0014 | 1.8650 ± 0.0019 | 0.0942 ± 0.0010 |
| SNGP | 0.01 | 1.7961 ± 0.0021 | 1.9032 ± 0.0021 | 1.7590 ± 0.0027 | 0.1531 ± 0.0012 |
| SNGP-MIL | 0.01 | 1.8760 ± 0.0025 | 1.8292 ± 0.0026 | 1.7213 ± 0.0034 | 0.1831 ± 0.0019 |
| SNGP-RE | 0.01 | 1.8629 ± 0.0019 | 1.9397 ± 0.0014 | 1.8431 ± 0.0024 | 0.1111 ± 0.0014 |
| SNGP-EAT | 0.01 | 1.7445 ± 0.0036 | 1.7739 ± 0.0037 | 1.6646 ± 0.0041 | 0.1387 ± 0.0015 |

Supplementary Table 26: **Racial fairness gaps of Inception-v3-based models in average set size on the CPTAC testing set with respect to three different error levels ($\alpha$).** The best-performing model for each setting is highlighted in bold. Results are presented as the mean ± 95% confidence intervals, derived from 20 random data splits and corresponding model training sessions, with each including 500 conformal prediction evaluations based on random partitions of the calibration and testing datasets.

| **Model** | $\alpha$ | **Average set size across sexual groups** | | |
|---|---|---|---|---|
| | | Female | Male | Fairness gap |
| Deterministic | 0.10 | 1.1509 ± 0.0020 | 1.1424 ± 0.0018 | 0.0667 ± 0.0011 |
| MC Dropout | 0.10 | 1.1588 ± 0.0021 | 1.1453 ± 0.0018 | 0.0676 ± 0.0012 |
| SNGP | 0.10 | 1.0644 ± 0.0015 | 1.0730 ± 0.0014 | 0.0445 ± 0.0008 |
| SNGP-MIL | 0.10 | 1.0070 ± 0.0013 | 1.0023 ± 0.0010 | **0.0322 ± 0.0006** |
| SNGP-RE | 0.10 | 1.2215 ± 0.0030 | 1.1715 ± 0.0022 | 0.0894 ± 0.0015 |
| SNGP-EAT | 0.10 | 1.0118 ± 0.0013 | 1.0120 ± 0.0011 | 0.0323 ± 0.0006 |
| Deterministic | 0.05 | 1.3772 ± 0.0029 | 1.3533 ± 0.0026 | 0.1011 ± 0.0015 |
| MC Dropout | 0.05 | 1.3774 ± 0.0027 | 1.3455 ± 0.0023 | 0.1031 ± 0.0015 |
| SNGP | 0.05 | 1.2445 ± 0.0023 | 1.2410 ± 0.0019 | 0.0797 ± 0.0012 |
| SNGP-MIL | 0.05 | 1.3153 ± 0.0050 | 1.1979 ± 0.0028 | 0.1432 ± 0.0026 |
| SNGP-RE | 0.05 | 1.4525 ± 0.0036 | 1.3933 ± 0.0031 | 0.1128 ± 0.0016 |
| SNGP-EAT | 0.05 | 1.1764 ± 0.0021 | 1.1676 ± 0.0021 | **0.0613 ± 0.0010** |
| Deterministic | 0.01 | 1.7011 ± 0.0035 | 1.6489 ± 0.0034 | 0.0978 ± 0.0014 |
| MC Dropout | 0.01 | 1.6920 ± 0.0036 | 1.6526 ± 0.0032 | 0.0888 ± 0.0013 |
| SNGP | 0.01 | 1.6168 ± 0.0037 | 1.5773 ± 0.0035 | 0.0872 ± 0.0013 |
| SNGP-MIL | 0.01 | 1.7462 ± 0.0037 | 1.6068 ± 0.0046 | 0.1671 ± 0.0024 |
| SNGP-RE | 0.01 | 1.7777 ± 0.0036 | 1.7584 ± 0.0037 | **0.0767 ± 0.0013** |
| SNGP-EAT | 0.01 | 1.6168 ± 0.0042 | 1.6480 ± 0.0044 | 0.0880 ± 0.0013 |

Supplementary Table 27: **Sexual fairness gaps of Inception-v3-based models in average set size on the TCGA testing set with respect to three different error levels ($\alpha$).** The best-performing model for each setting is highlighted in bold. Results are presented as the mean ± 95% confidence intervals, derived from 20 random data splits and corresponding model training sessions, with each including 500 conformal prediction evaluations based on random partitions of the calibration and testing datasets.

| **Model** | $\alpha$ | **Average set size across sexual groups** | | |
|---|---|---|---|---|
| | | Female | Male | Fairness gap |
| Deterministic | 0.10 | 1.5483 ± 0.0035 | 1.4497 ± 0.0027 | 0.1098 ± 0.0012 |
| MC Dropout | 0.10 | 1.5188 ± 0.0035 | 1.4242 ± 0.0028 | 0.0994 ± 0.0012 |
| SNGP | 0.10 | 1.3279 ± 0.0033 | 1.2893 ± 0.0024 | 0.0606 ± 0.0009 |
| SNGP-MIL | 0.10 | 1.2671 ± 0.0049 | 1.2149 ± 0.0034 | 0.0735 ± 0.0014 |
| SNGP-RE | 0.10 | 1.5405 ± 0.0036 | 1.4667 ± 0.0029 | 0.0980 ± 0.0012 |
| SNGP-EAT | 0.10 | 1.1277 ± 0.0021 | 1.1327 ± 0.0021 | **0.0296 ± 0.0005** |
| Deterministic | 0.05 | 1.7508 ± 0.0024 | 1.6562 ± 0.0024 | 0.1002 ± 0.0010 |
| MC Dropout | 0.05 | 1.7382 ± 0.0025 | 1.6414 ± 0.0025 | 0.0980 ± 0.0010 |
| SNGP | 0.05 | 1.5687 ± 0.0032 | 1.4942 ± 0.0025 | 0.0881 ± 0.0010 |
| SNGP-MIL | 0.05 | 1.5438 ± 0.0047 | 1.4479 ± 0.0033 | 0.1191 ± 0.0013 |
| SNGP-RE | 0.05 | 1.7060 ± 0.0029 | 1.6515 ± 0.0027 | 0.0786 ± 0.0010 |
| SNGP-EAT | 0.05 | 1.3076 ± 0.0031 | 1.3169 ± 0.0026 | **0.0451 ± 0.0006** |
| Deterministic | 0.01 | 1.9135 ± 0.0012 | 1.8721 ± 0.0017 | 0.0463 ± 0.0008 |
| MC Dropout | 0.01 | 1.9120 ± 0.0012 | 1.8682 ± 0.0017 | 0.0464 ± 0.0008 |
| SNGP | 0.01 | 1.8179 ± 0.0022 | 1.7733 ± 0.0025 | 0.0525 ± 0.0008 |
| SNGP-MIL | 0.01 | 1.8249 ± 0.0027 | 1.7599 ± 0.0030 | 0.0902 ± 0.0011 |
| SNGP-RE | 0.01 | 1.8784 ± 0.0019 | 1.8519 ± 0.0022 | **0.0368 ± 0.0007** |
| SNGP-EAT | 0.01 | 1.6981 ± 0.0041 | 1.7014 ± 0.0038 | 0.0488 ± 0.0007 |

Supplementary Table 28: **Sexual fairness gaps of Inception-v3-based models in average set size on the CPTAC testing set with respect to three different error levels ($\alpha$).** The best-performing model for each setting is highlighted in bold. Results are presented as the mean ± 95% confidence intervals, derived from 20 random data splits and corresponding model training sessions, with each including 500 conformal prediction evaluations based on random partitions of the calibration and testing datasets.

| **Model** | **Evaluation dataset** | $c$ | **Accuracy** | **AUROC** |
|---|---|---|---|---|
| UNI | TCGA | None | 0.9313 ± 0.0060 | **0.9747 ± 0.0034** |
| UNI-SNGP | TCGA | 2.0 | 0.9377 ± 0.0003 | 0.9734 ± 0.0001 |
| UNI-SNGP-EAT | TCGA | 2.0 | **0.9382 ± 0.0003** | 0.9737 ± 0.0001 |
| CONCH | TCGA | None | 0.9210 ± 0.0077 | 0.9729 ± 0.0041 |
| CONCH-SNGP | TCGA | 2.0 | 0.9224 ± 0.0089 | 0.9747 ± 0.0038 |
| CONCH-SNGP-EAT | TCGA | 2.0 | **0.9277 ± 0.0003** | **0.9762 ± 0.0001** |
| Prov-GigaPath | TCGA | None | 0.9310 ± 0.0065 | 0.9791 ± 0.0028 |
| Prov-GigaPath-EAT | TCGA | None | **0.9342 ± 0.0072** | **0.9797 ± 0.0029** |
| TITAN | TCGA | None | 0.9329 ± 0.0064 | **0.9767 ± 0.0028** |
| TITAN-EAT | TCGA | None | **0.9376 ± 0.0056** | 0.9766 ± 0.0030 |
| UNI | CPTAC | None | 0.9464 ± 0.0028 | 0.9753 ± 0.0006 |
| UNI-SNGP | CPTAC | 2.0 | 0.9458 ± 0.0038 | 0.9774 ± 0.0013 |
| UNI-SNGP-EAT | CPTAC | 2.0 | **0.9470 ± 0.0001** | **0.9786 ± 0.0001** |
| CONCH | CPTAC | None | 0.9470 ± 0.0028 | 0.9732 ± 0.0041 |
| CONCH-SNGP | CPTAC | 2.0 | 0.9476 ± 0.0028 | **0.9836 ± 0.0013** |
| CONCH-SNGP-EAT | CPTAC | 2.0 | **0.9524 ± 0.0000** | 0.9821 ± 0.0001 |
| Prov-GigaPath | CPTAC | None | **0.9411 ± 0.0078** | 0.9789 ± 0.0026 |
| Prov-GigaPath-EAT | CPTAC | None | **0.9411 ± 0.0072** | **0.9800 ± 0.0025** |
| TITAN | CPTAC | None | **0.9643 ± 0.0000** | 0.9747 ± 0.0000 |
| TITAN-EAT | CPTAC | None | **0.9643 ± 0.0000** | **0.9765 ± 0.0004** |

Supplementary Table 29: **Main classification results of foundation models**. For Prov-GigaPath and TITAN, we do not enforce spectral normalization due to the following reasons: 1) Prov-GigaPath employs a two-stage pretraining strategy, and enforcing spectral normalization in the slide encoder (LongNet) could negatively impact the pretrained weights; 2) TITAN leverages linear probing on the slide embeddings without further fine-tuning the slide encoder, making the impose of spectral normalization unnecessary. These observations suggest that TRUECAM is well modularized and its modules can be effectively integrated into various models. For simplicity, we set the mask tile threshold to 0.4 for all models in TCGA and CPTAC. The exception is the CPTAC cohort of Prov-GigaPath and TITAN, where the threshold is set to 0.8 in our experiments. More results on ablation study of mask tile threshold can be seen in Supplementary Table 43 - 46. Note that TITAN-preview is abbreviated as TITAN throughout this paper.

| Model | Evaluation dataset | $c$ | Accuracy | AUROC |
|---|---|---|---|---|
| CONCH | TCGA | None | 0.9210 ± 0.0079 | 0.9729 ± 0.0042 |
| CONCH-SNGP | TCGA | 0.5 | 0.9002 ± 0.0069 | 0.9602 ± 0.0043 |
| CONCH-SNGP | TCGA | 1.0 | **0.9247 ± 0.0074** | 0.9746 ± 0.0041 |
| CONCH-SNGP | TCGA | 2.0 | 0.9224 ± 0.0089 | **0.9747 ± 0.0038** |
| CONCH-SNGP | TCGA | 3.0 | 0.9224 ± 0.0097 | 0.9727 ± 0.0059 |
| CONCH-SNGP | TCGA | 4.0 | 0.9222 ± 0.0091 | 0.9743 ± 0.0041 |
| CONCH-SNGP | TCGA | 5.0 | 0.9222 ± 0.0091 | 0.9743 ± 0.0041 |
| CONCH-SNGP | TCGA | 6.0 | 0.9222 ± 0.0091 | 0.9743 ± 0.0041 |

Supplementary Table 30: **Impact of spectral normalization factor $c$ on the performance of CONCH and CONCH-SNGP on TCGA (n=941)**. See Methods in the paper for more details. Tile representations extracted from CONCH, CONCH and CONCH-SNGP were trained using the ABMIL framework and evaluated on a curated train-validation-test with a split ratio of 0.65:0.15:0.20, consisting of 610, 142, and 189 patients, respectively. The experimental setting was identical with that used in the development of Inception models. The model with the best validation accuracy was saved for performance evaluation. Note that certain spectral normalization factors ($\hat{\lambda} = 5.0$, $\hat{\lambda} = 6.0$) yield the same performance in terms of accuracy and AUROC. This phenomenon is attributed to the fact that higher spectral normalization factor imposes a loose constraint on the neural network weights. In other words, a larger $c$ enforces a loose constraint on the distance-preserving data transformation. In such cases, the performance is primarily influenced by the Gaussian layer rather than the spectral normalization factor. Best-performing model for each case is highlighted in bold, and results are reported as mean ± 95% confidence interval (CI) from 20 experimental replicates.

| Model | Evaluation dataset | $c$ | Accuracy | AUROC |
|---|---|---|---|---|
| CONCH | CPTAC | None | 0.9470 ± 0.0029 | 0.9732 ± 0.0042 |
| CONCH-SNGP | CPTAC | 0.5 | 0.9167 ± 0.0000 | 0.9725 ± 0.0003 |
| CONCH-SNGP | CPTAC | 1.0 | 0.9405 ± 0.0000 | 0.9835 ± 0.0003 |
| CONCH-SNGP | CPTAC | 2.0 | **0.9476 ± 0.0028** | **0.9836 ± 0.0013** |
| CONCH-SNGP | CPTAC | 3.0 | 0.9470 ± 0.0029 | 0.9718 ± 0.0051 |
| CONCH-SNGP | CPTAC | 4.0 | 0.9470 ± 0.0029 | 0.9730 ± 0.0042 |
| CONCH-SNGP | CPTAC | 5.0 | 0.9464 ± 0.0029 | 0.9737 ± 0.0043 |
| CONCH-SNGP | CPTAC | 6.0 | 0.9464 ± 0.0029 | 0.9737 ± 0.0043 |

Supplementary Table 31: **Impact of $c$ on the performance of CONCH and CONCH-SNGP with respect to the CPTAC cohort (n=416).** Tile representations extracted from CONCH, CONCH and CONCH-SNGP were trained using the ABMIL framework and evaluated on a curated train-validation-test with a split ratio of 0.65:0.15:0.20, consisting of 270, 62, and 84 patients, respectively. Best-performing model for each case is highlighted in bold, and results are reported as mean ± 95% confidence interval (CI) from 20 experimental replicates.

| Model | Evaluation dataset | $c$ | Empirical coverage | |
|---|---|---|---|---|
| | | | $\alpha = 0.05$ | $\alpha = 0.01$ |
| CONCH | TCGA | None | 0.9559 ± 0.0006 | 0.9900 ± 0.0003 |
| CONCH-SNGP | TCGA | 0.5 | 0.9560 ± 0.0006 | 0.9899 ± 0.0003 |
| CONCH-SNGP | TCGA | 1.0 | 0.9557 ± 0.0006 | 0.9899 ± 0.0003 |
| CONCH-SNGP | TCGA | 2.0 | 0.9563 ± 0.0006 | 0.9900 ± 0.0003 |
| CONCH-SNGP | TCGA | 3.0 | 0.9560 ± 0.0006 | 0.9900 ± 0.0003 |
| CONCH-SNGP | TCGA | 4.0 | 0.9560 ± 0.0006 | 0.9900 ± 0.0003 |
| CONCH-SNGP | TCGA | 5.0 | 0.9560 ± 0.0006 | 0.9900 ± 0.0003 |
| CONCH-SNGP | TCGA | 6.0 | 0.9560 ± 0.0006 | 0.9900 ± 0.0003 |

Supplementary Table 32: **Impact of $c$ on patient-level empirical coverage of conformalized CONCH and CONCH-SNGP with respect to TCGA test data at various miscoverage rates ($\alpha = 0.05$, $\alpha = 0.01$).** Note that we do not report the empirical coverage at $\alpha = 0.10$ because the base CONCH model has a miscoverage rate less than 0.1. Results are presented with mean ± 95% CI from 20 experimental replicates with each using 500 randomly selected calibration/test permutations.

| Model | Evaluation dataset | $c$ | Empirical coverage | |
|---|---|---|---|---|
| | | | $\alpha = 0.05$ | $\alpha = 0.01$ |
| CONCH | CPTAC | None | 0.9554 ± 0.0006 | 0.9890 ± 0.0003 |
| CONCH-SNGP | CPTAC | 0.5 | 0.9539 ± 0.0006 | 0.9891 ± 0.0003 |
| CONCH-SNGP | CPTAC | 1.0 | 0.9543 ± 0.0006 | 0.9891 ± 0.0003 |
| CONCH-SNGP | CPTAC | 2.0 | 0.9550 ± 0.0006 | 0.9890 ± 0.0003 |
| CONCH-SNGP | CPTAC | 3.0 | 0.9549 ± 0.0006 | 0.9890 ± 0.0003 |
| CONCH-SNGP | CPTAC | 4.0 | 0.9551 ± 0.0006 | 0.9890 ± 0.0003 |
| CONCH-SNGP | CPTAC | 5.0 | 0.9551 ± 0.0006 | 0.9890 ± 0.0003 |
| CONCH-SNGP | CPTAC | 6.0 | 0.9551 ± 0.0006 | 0.9890 ± 0.0003 |

Supplementary Table 33: **Impact of $c$ on patient-level empirical coverage of conformalized CONCH foundation models with respect to the CPTAC test data at various miscoverage rates ($\alpha = 0.05$, $\alpha = 0.01$).** Results are presented with mean ± 95% CI from 20 experimental replicates with each using 500 randomly selected calibration/test permutations.

| Model | Evaluation dataset | $c$ | Average set size | |
|---|---|---|---|---|
| | | | $\alpha = 0.05$ | $\alpha = 0.01$ |
| CONCH | TCGA | None | 1.1159 ± 0.0016 | 1.5478 ± 0.0051 |
| CONCH-SNGP | TCGA | 0.5 | 1.1730 ± 0.0018 | 1.5993 ± 0.0036 |
| CONCH-SNGP | TCGA | 1.0 | **1.0982 ± 0.0014** | **1.5088 ± 0.0050** |
| CONCH-SNGP | TCGA | 2.0 | 1.1068 ± 0.0016 | **1.5088 ± 0.0048** |
| CONCH-SNGP | TCGA | 3.0 | 1.1120 ± 0.0018 | 1.5406 ± 0.0053 |
| CONCH-SNGP | TCGA | 4.0 | 1.1111 ± 0.0017 | 1.5248 ± 0.0050 |
| CONCH-SNGP | TCGA | 5.0 | 1.1111 ± 0.0017 | 1.5248 ± 0.0050 |
| CONCH-SNGP | TCGA | 6.0 | 1.1111 ± 0.0017 | 1.5248 ± 0.0050 |

Supplementary Table 34: **Impact of $c$ on patient-level average set size of conformalized CONCH and CONCH-SNGP with respect to the TCGA test data at various miscoverage rates ($\alpha = 0.05$, $\alpha = 0.01$).** Results are presented with mean ± 95% CI from 20 experimental replicates with each using 500 randomly selected calibration/test permutations.

| Model | Evaluation dataset | $c$ | Average set size | |
|---|---|---|---|---|
| | | | $\alpha = 0.05$ | $\alpha = 0.01$ |
| CONCH | CPTAC | None | 1.0292 ± 0.0012 | 1.6615 ± 0.0045 |
| CONCH-SNGP | CPTAC | 0.5 | 1.1000 ± 0.0010 | **1.5054 ± 0.0032** |
| CONCH-SNGP | CPTAC | 1.0 | **1.0131 ± 0.0008** | 1.5916 ± 0.0049 |
| CONCH-SNGP | CPTAC | 2.0 | 1.0174 ± 0.0008 | 1.5156 ± 0.0045 |
| CONCH-SNGP | CPTAC | 3.0 | 1.0240 ± 0.0011 | 1.6385 ± 0.0045 |
| CONCH-SNGP | CPTAC | 4.0 | 1.0246 ± 0.0011 | 1.6380 ± 0.0043 |
| CONCH-SNGP | CPTAC | 5.0 | 1.0252 ± 0.0011 | 1.6376 ± 0.0044 |
| CONCH-SNGP | CPTAC | 6.0 | 1.0252 ± 0.0011 | 1.6376 ± 0.0044 |

Supplementary Table 35: **Impact of $c$ on patient-level average set size of conformalized CONCH and CONCH-SNGP with respect to the CPTAC test data at various miscoverage rates ($\alpha = 0.05$, $\alpha = 0.01$).** Results are presented with mean ± 95% CI from 20 experimental replicates with each using 500 randomly selected calibration/test permutations.

| Model | Evaluation dataset | $c$ | Definitive-answer error rate | | |
|---|---|---|---|---|---|
| | | | W/O CP | $\alpha = 0.05$ | $\alpha = 0.01$ |
| CONCH | TCGA | None | 0.0790 ± 0.0079 | 0.0470 ± 0.0005 | 0.0165 ± 0.0005 |
| CONCH-SNGP | TCGA | 0.5 | 0.0998 ± 0.0069 | 0.0509 ± 0.0006 | 0.0192 ± 0.0005 |
| CONCH-SNGP | TCGA | 1.0 | **0.0753 ± 0.0074** | 0.0468 ± 0.0005 | 0.0146 ± 0.0004 |
| CONCH-SNGP | TCGA | 2.0 | 0.0776 ± 0.0089 | **0.0462 ± 0.0005** | **0.0145 ± 0.0004** |
| CONCH-SNGP | TCGA | 3.0 | 0.0776 ± 0.0097 | 0.0467 ± 0.0005 | 0.0167 ± 0.0005 |
| CONCH-SNGP | TCGA | 4.0 | 0.0778 ± 0.0091 | 0.0467 ± 0.0005 | 0.0151 ± 0.0005 |
| CONCH-SNGP | TCGA | 5.0 | 0.0778 ± 0.0091 | 0.0467 ± 0.0005 | 0.0151 ± 0.0005 |
| CONCH-SNGP | TCGA | 6.0 | 0.0778 ± 0.0091 | 0.0467 ± 0.0005 | 0.0151 ± 0.0005 |

Supplementary Table 36: **Impact of $c$ on patient-level definitive-answer error rate of CONCH and CONCH-SNGP with respect to the TCGA test data before and after applying conformal prediction**. Best-performing model for each case is highlighted in bold, and results are presented with mean ± 95% CI from 20 experimental replicates with each using 500 randomly selected calibration/test permutations (except W/O CP error rate).

| Model | Evaluation dataset | $c$ | Definitive-answer error rate | | |
|---|---|---|---|---|---|
| | | | W/O CP | $\alpha = 0.05$ | $\alpha = 0.01$ |
| CONCH | CPTAC | None | 0.0530 ± 0.0029 | 0.0390 ± 0.0004 | 0.0276 ± 0.0007 |
| CONCH-SNGP | CPTAC | 0.5 | 0.0833 ± 0.0000 | 0.0498 ± 0.0006 | **0.0168 ± 0.0004** |
| CONCH-SNGP | CPTAC | 1.0 | 0.0595 ± 0.0000 | 0.0394 ± 0.0004 | 0.0191 ± 0.0004 |
| CONCH-SNGP | CPTAC | 2.0 | **0.0524 ± 0.0028** | 0.0392 ± 0.0004 | 0.0170 ± 0.0004 |
| CONCH-SNGP | CPTAC | 3.0 | 0.0530 ± 0.0029 | 0.0391 ± 0.0004 | 0.0254 ± 0.0007 |
| CONCH-SNGP | CPTAC | 4.0 | 0.0530 ± 0.0029 | **0.0389 ± 0.0004** | 0.0241 ± 0.0006 |
| CONCH-SNGP | CPTAC | 5.0 | 0.0536 ± 0.0029 | **0.0389 ± 0.0004** | 0.0240 ± 0.0006 |
| CONCH-SNGP | CPTAC | 6.0 | 0.0536 ± 0.0029 | **0.0389 ± 0.0004** | 0.0240 ± 0.0006 |

Supplementary Table 37: **Impact of $c$ on patient-level definitive-answer error rate of CONCH and CONCH-SNGP with respect to the CPTAC test data before and after applying conformal prediction**. Best-performing model for each case is highlighted in bold, and results are presented with mean ± 95% CI from 20 experimental replicates with each using 500 randomly selected calibration/test permutations (except W/O CP definitive-answer error rate).

| Model | Evaluation dataset | $c$ | Definitive-answer error rate | | |
|---|---|---|---|---|---|
| | | | W/O CP | $\alpha = 0.05$ | $\alpha = 0.01$ |
| CONCH | TCGA | None | 0.9210 ± 0.0079 | 0.8386 ± 0.0013 | 0.4421 ± 0.0050 |
| CONCH-SNGP | TCGA | 0.5 | 0.9002 ± 0.0069 | 0.7830 ± 0.0015 | 0.3906 ± 0.0034 |
| CONCH-SNGP | TCGA | 1.0 | **0.9247 ± 0.0074** | **0.8563 ± 0.0011** | **0.4812 ± 0.0048** |
| CONCH-SNGP | TCGA | 2.0 | 0.9224 ± 0.0089 | 0.8481 ± 0.0013 | **0.4812 ± 0.0046** |
| CONCH-SNGP | TCGA | 3.0 | 0.9224 ± 0.0097 | 0.8424 ± 0.0015 | 0.4494 ± 0.0051 |
| CONCH-SNGP | TCGA | 4.0 | 0.9222 ± 0.0091 | 0.8433 ± 0.0014 | 0.4652 ± 0.0049 |
| CONCH-SNGP | TCGA | 5.0 | 0.9222 ± 0.0091 | 0.8433 ± 0.0014 | 0.4652 ± 0.0049 |
| CONCH-SNGP | TCGA | 6.0 | 0.9222 ± 0.0091 | 0.8433 ± 0.0014 | 0.4652 ± 0.0049 |

Supplementary Table 38: **Impact of $c$ on patient-level Definitive-answer recall rate of CONCH foundation models with respect to the TCGA test data before and after applying conformal prediction**. Best-performing model for each case is highlighted in bold, and results are presented with mean ± 95% CI from 20 experimental replicates with each using 500 randomly selected calibration/test permutations (except W/O CP definitive-answer error rate).

| Model | Evaluation dataset | $c$ | Definitive-answer error rate | | |
|---|---|---|---|---|---|
| | | | W/O CP | $\alpha = 0.05$ | $\alpha = 0.01$ |
| CONCH | CPTAC | None | 0.9470 ± 0.0029 | 0.9196 ± 0.0009 | 0.3274 ± 0.0042 |
| CONCH-SNGP | CPTAC | 0.5 | 0.9167 ± 0.0000 | 0.8538 ± 0.0006 | 0.4837 ± 0.0029 |
| CONCH-SNGP | CPTAC | 1.0 | 0.9405 ± 0.0000 | **0.9344 ± 0.0005** | 0.3974 ± 0.0046 |
| CONCH-SNGP | CPTAC | 2.0 | **0.9476 ± 0.0028** | 0.9309 ± 0.0004 | **0.4733 ± 0.0042** |
| CONCH-SNGP | CPTAC | 3.0 | 0.9470 ± 0.0029 | 0.9241 ± 0.0007 | 0.3503 ± 0.0042 |
| CONCH-SNGP | CPTAC | 4.0 | 0.9470 ± 0.0029 | 0.9238 ± 0.0007 | 0.3510 ± 0.0041 |
| CONCH-SNGP | CPTAC | 5.0 | 0.9464 ± 0.0029 | 0.9231 ± 0.0007 | 0.3513 ± 0.0041 |
| CONCH-SNGP | CPTAC | 6.0 | 0.9464 ± 0.0029 | 0.9231 ± 0.0007 | 0.3513 ± 0.0041 |

Supplementary Table 39: **Impact of $c$ on patient-level definitive-answer recall rate of CONCH and CONCH-SNGP with respect to the CPTAC test data before and after applying conformal prediction**. Best-performing model for each case is highlighted in bold, and results are presented with mean ± 95% CI from 20 experimental replicates with each using 500 randomly selected calibration/test permutations (except W/O CP error rate).

| | Uncertainty-based OOD score | | Probability-based OOD score | |
|---|---|---|---|---|
| | AUROC | AUPR | AUROC | AUPR |
| UNI | - | - | 0.8708 ± 0.0132 | 0.9385 ± 0.0065 |
| UNI-SNGP | 0.9639 ± 0.0051 | 0.9850 ± 0.0031 | 0.8842 ± 0.0290 | 0.9444 ± 0.0136 |
| UNI-SNGP-EAT | **0.9671 ± 0.0045** | **0.9858 ± 0.0029** | 0.8824 ± 0.0332 | 0.9431 ± 0.0152 |
| CONCH | - | - | 0.7307 ± 0.0306 | 0.8879 ± 0.0147 |
| CONCH-SNGP | **0.9840 ± 0.0031** | **0.9931 ± 0.0016** | 0.7477 ± 0.0251 | 0.8912 ± 0.0129 |
| CONCH-SNGP-EAT | 0.9836 ± 0.0031 | 0.9928 ± 0.0017 | 0.7495 ± 0.0275 | 0.9007 ± 0.0127 |
| Prov-GigaPath | - | - | 0.7256 ± 0.0245 | 0.8903 ± 0.0104 |
| Prov-GigaPath-EAT | **0.9788 ± 0.0073** | **0.9916 ± 0.0032** | 0.7314 ± 0.0224 | 0.8948 ± 0.0091 |
| TITAN | - | - | 0.7427 ± 0.0371 | 0.8938 ± 0.0159 |
| TITAN-EAT | **0.9906 ± 0.0018** | **0.9961 ± 0.0010** | 0.8312 ± 0.0148 | 0.9268 ± 0.0066 |

Supplementary Table 40: **Patient-level OOD detection performance of UNI, CONCH, Prov-GigaPath and TITAN, where the TCGA test set (n=189) serves as the in-domain dataset, and the concatenated UVM (n=70), UCS (n=57), ACC (n=92), BLCA (n=412) serves as OOD datasets**. We treat the label of in-domain test data as 0 and the label of OOD data as 1 to derive the AUROC and AUPR. Best-performing model for each case is highlighted in bold, and results are presented with mean ± 95% CI from 20 experimental replicates.

| **Model** | **In-D** | $c$ | **AUROC** | | |
|---|---|---|---|---|---|
| | | | UCS | UVM | BLCA |
| CONCH-SNGP | TCGA | 0.5 | 0.9704 ± 0.0098 | 0.9899 ± 0.0031 | 0.9216 ± 0.0159 |
| CONCH-SNGP | TCGA | 1.0 | **0.9905 ± 0.0020** | 0.9968 ± 0.0008 | 0.9760 ± 0.0022 |
| CONCH-SNGP | TCGA | 2.0 | 0.9903 ± 0.0025 | **0.9970 ± 0.0013** | 0.9786 ± 0.0040 |
| CONCH-SNGP | TCGA | 3.0 | 0.9896 ± 0.0031 | 0.9962 ± 0.0019 | 0.9785 ± 0.0041 |
| CONCH-SNGP | TCGA | 4.0 | 0.9900 ± 0.0026 | 0.9968 ± 0.0011 | **0.9789 ± 0.0033** |
| CONCH-SNGP | TCGA | 5.0 | 0.9900 ± 0.0026 | 0.9968 ± 0.0011 | **0.9789 ± 0.0033** |
| CONCH-SNGP | TCGA | 6.0 | 0.9900 ± 0.0026 | 0.9968 ± 0.0011 | **0.9789 ± 0.0033** |

| **Model** | **In-D** | $c$ | **AUROC** | |
|---|---|---|---|---|
| | | | ACC | Average |
| CONCH-SNGP | TCGA | 0.5 | 0.9717 ± 0.0064 | 0.9409 ± 0.0122 |
| CONCH-SNGP | TCGA | 1.0 | 0.9922 ± 0.0018 | 0.9820 ± 0.0018 |
| CONCH-SNGP | TCGA | 2.0 | 0.9946 ± 0.0021 | 0.9840 ± 0.0031 |
| CONCH-SNGP | TCGA | 3.0 | 0.9942 ± 0.0029 | 0.9837 ± 0.0034 |
| CONCH-SNGP | TCGA | 4.0 | **0.9951 ± 0.0017** | **0.9843 ± 0.0026** |
| CONCH-SNGP | TCGA | 5.0 | **0.9951 ± 0.0017** | **0.9843 ± 0.0026** |
| CONCH-SNGP | TCGA | 6.0 | **0.9951 ± 0.0017** | **0.9843 ± 0.0026** |

Supplementary Table 41: **Patient-level fine-grained OOD detection performance of CONCH-SNGP in terms of AUROC and AUPR with In-D TCGA using uncertainty-based OOD score**. Impact of spectral normalization factor $c$ on the performance of CONCH-SNGP is reported. Best-performing model for each case is highlighted in bold, and results are presented with mean ± 95% CI from 20 experimental replicates.

| **Model** | **In-D** | $c$ | **AUROC** | | |
|---|---|---|---|---|---|
| | | | UCS | UVM | BLCA |
| CONCH-SNGP | CPTAC | 0.5 | 0.8985 ± 0.0174 | 0.9581 ± 0.0073 | 0.8377 ± 0.0192 |
| CONCH-SNGP | CPTAC | 1.0 | **0.9939 ± 0.0016** | **0.9998 ± 0.0001** | **0.9825 ± 0.0033** |
| CONCH-SNGP | CPTAC | 2.0 | 0.9713 ± 0.0036 | 0.9980 ± 0.0004 | 0.9363 ± 0.0049 |
| CONCH-SNGP | CPTAC | 3.0 | 0.9754 ± 0.0033 | 0.9990 ± 0.0005 | 0.9470 ± 0.0049 |
| CONCH-SNGP | CPTAC | 4.0 | 0.9761 ± 0.0024 | 0.9990 ± 0.0005 | 0.9469 ± 0.0049 |
| CONCH-SNGP | CPTAC | 5.0 | 0.9749 ± 0.0026 | 0.9988 ± 0.0005 | 0.9451 ± 0.0047 |
| CONCH-SNGP | CPTAC | 6.0 | 0.9749 ± 0.0026 | 0.9988 ± 0.0005 | 0.9451 ± 0.0047 |

| **Model** | **In-D** | $c$ | **AUROC** | |
|---|---|---|---|---|
| | | | ACC | Average |
| CONCH-SNGP | CPTAC | 0.5 | 0.9475 ± 0.0098 | 0.8726 ± 0.0161 |
| CONCH-SNGP | CPTAC | 1.0 | **0.9980 ± 0.0004** | **0.9877 ± 0.0023** |
| CONCH-SNGP | CPTAC | 2.0 | 0.9921 ± 0.0010 | 0.9544 ± 0.0036 |
| CONCH-SNGP | CPTAC | 3.0 | 0.9941 ± 0.0013 | 0.9622 ± 0.0037 |
| CONCH-SNGP | CPTAC | 4.0 | 0.9942 ± 0.0015 | 0.9622 ± 0.0036 |
| CONCH-SNGP | CPTAC | 5.0 | 0.9938 ± 0.0015 | 0.9608 ± 0.0035 |
| CONCH-SNGP | CPTAC | 6.0 | 0.9938 ± 0.0015 | 0.9608 ± 0.0035 |

Supplementary Table 42: **Patient-level fine-grained OOD detection performance of CONCH in terms of AUROC and AUPR with In-D CPTAC using uncertainty-based OOD score**. Impact of spectral normalization factor $c$ on the performance of CONCH-SNGP is reported. Best-performing model for each case is highlighted in bold, and results are presented with mean ± 95% CI from 20 experimental replicates.

| Mask Ratio | Evaluation dataset | Random | | Ambiguity | |
|---|---|---|---|---|---|
| | | Accuracy | AUROC | Accuracy | AUROC |
| 0.001 | TCGA | 0.8348 ± 0.0154 | 0.9366 ± 0.0071 | 0.9340 ± 0.0077 | 0.9681 ± 0.0040 |
| 0.01 | TCGA | 0.9145 ± 0.0091 | 0.9702 ± 0.0038 | **0.9411 ± 0.0070** | 0.9711 ± 0.0039 |
| 0.1 | TCGA | 0.9335 ± 0.0080 | 0.9730 ± 0.0034 | 0.9401 ± 0.0069 | 0.9731 ± 0.0034 |
| 0.2 | TCGA | 0.9369 ± 0.0075 | 0.9734 ± 0.0031 | 0.9401 ± 0.0067 | 0.9734 ± 0.0033 |
| 0.3 | TCGA | 0.9348 ± 0.0074 | 0.9734 ± 0.0032 | 0.9393 ± 0.0066 | 0.9737 ± 0.0032 |
| 0.4 | TCGA | 0.9356 ± 0.0073 | 0.9734 ± 0.0031 | 0.9382 ± 0.0067 | 0.9737 ± 0.0032 |
| 0.5 | TCGA | 0.9356 ± 0.0069 | 0.9735 ± 0.0031 | 0.9382 ± 0.0066 | 0.9737 ± 0.0032 |
| 0.6 | TCGA | 0.9363 ± 0.0071 | 0.9732 ± 0.0032 | 0.9382 ± 0.0066 | **0.9738 ± 0.0032** |
| 0.7 | TCGA | 0.9371 ± 0.0072 | 0.9731 ± 0.0032 | 0.9382 ± 0.0068 | **0.9738 ± 0.0032** |
| 0.8 | TCGA | 0.9371 ± 0.0066 | 0.9732 ± 0.0032 | 0.9379 ± 0.0066 | 0.9737 ± 0.0032 |
| 0.9 | TCGA | 0.9374 ± 0.0070 | 0.9734 ± 0.0031 | 0.9382 ± 0.0068 | 0.9737 ± 0.0032 |
| No mask | TCGA | 0.9377 ± 0.0069 | 0.9734 ± 0.0032 | 0.9377 ± 0.0069 | 0.9734 ± 0.0032 |

Supplementary Table 43: **Impact of mask ratio and elimination strategies on patient-level accuracy and AUROC of UNI-SNGP (no retraining) with respect to the TCGA test data**. In the following supplementary material, unless otherwise specified, the spectral normalization factor $c$ of UNI-SNGP is fixed at 2.0. Best-performing model for each case is highlighted in bold, and results are presented with mean ± 95% CI from 20 experimental replicates.

| Mask Ratio | Evaluation dataset | Random | | Ambiguity | |
|---|---|---|---|---|---|
| | | Accuracy | AUROC | Accuracy | AUROC |
| 0.001 | CPTAC | 0.8613 ± 0.0134 | 0.9492 ± 0.0058 | 0.8833 ± 0.0088 | 0.9706 ± 0.0027 |
| 0.01 | CPTAC | 0.9125 ± 0.0081 | 0.9741 ± 0.0022 | 0.9161 ± 0.0064 | 0.9761 ± 0.0029 |
| 0.1 | CPTAC | 0.9375 ± 0.0044 | 0.9768 ± 0.0014 | 0.9381 ± 0.0064 | 0.9783 ± 0.0017 |
| 0.2 | CPTAC | 0.9393 ± 0.0044 | 0.9771 ± 0.0013 | 0.9429 ± 0.0053 | 0.9782 ± 0.0016 |
| 0.3 | CPTAC | 0.9435 ± 0.0036 | 0.9774 ± 0.0011 | 0.9435 ± 0.0040 | **0.9786 ± 0.0015** |
| 0.4 | CPTAC | 0.9446 ± 0.0033 | 0.9773 ± 0.0012 | **0.9470 ± 0.0034** | **0.9786 ± 0.0016** |
| 0.5 | CPTAC | 0.9458 ± 0.0038 | 0.9775 ± 0.0012 | **0.9470 ± 0.0034** | **0.9786 ± 0.0015** |
| 0.6 | CPTAC | 0.9458 ± 0.0038 | 0.9776 ± 0.0012 | 0.9464 ± 0.0034 | **0.9786 ± 0.0015** |
| 0.7 | CPTAC | 0.9458 ± 0.0038 | 0.9775 ± 0.0013 | 0.9464 ± 0.0034 | 0.9783 ± 0.0015 |
| 0.8 | CPTAC | 0.9464 ± 0.0038 | 0.9775 ± 0.0014 | 0.9458 ± 0.0038 | 0.9782 ± 0.0015 |
| 0.9 | CPTAC | 0.9464 ± 0.0038 | 0.9775 ± 0.0013 | 0.9458 ± 0.0038 | 0.9779 ± 0.0015 |
| No mask | CPTAC | 0.9458 ± 0.0038 | 0.9774 ± 0.0013 | 0.9458 ± 0.0038 | 0.9774 ± 0.0013 |

Supplementary Table 44: **Impact of mask ratio and elimination strategies on patient-level accuracy and AUROC of UNI-SNGP (no retraining) with respect to the CPTAC test data**. Tile-level ambiguity predictor (AutoGluon) was trained and evaluated following a train-validation-test split ratio of 0.65:0.15:0.20, comprising 270, 62, and 84 patients, respectively. In the following supplementary material, unless otherwise specified, the evaluation of eliminating ambiguous tiles with UNI-SNGP and CONCH-SNGP of CPTAC are conducted on the test set (n=84). Best-performing model for each case is highlighted in bold, and results are presented with mean ± 95% CI from 20 experimental replicates.

| Mask Ratio | Evaluation dataset | **Random** | | **Ambiguity** | |
|---|---|---|---|---|---|
| | | Accuracy | AUROC | Accuracy | AUROC |
| 0.001 | TCGA | 0.8171 ± 0.0139 | 0.9064 ± 0.0091 | **0.9335 ± 0.0080** | 0.9704 ± 0.0054 |
| 0.01 | TCGA | 0.9068 ± 0.0075 | 0.9642 ± 0.0047 | **0.9335 ± 0.0072** | 0.9739 ± 0.0046 |
| 0.1 | TCGA | 0.9203 ± 0.0088 | 0.9729 ± 0.0042 | 0.9306 ± 0.0073 | 0.9751 ± 0.0040 |
| 0.2 | TCGA | 0.9211 ± 0.0095 | 0.9740 ± 0.0042 | 0.9295 ± 0.0074 | 0.9758 ± 0.0038 |
| 0.3 | TCGA | 0.9203 ± 0.0082 | 0.9739 ± 0.0041 | 0.9285 ± 0.0085 | 0.9760 ± 0.0037 |
| 0.4 | TCGA | 0.9216 ± 0.0098 | 0.9741 ± 0.0041 | 0.9277 ± 0.0079 | 0.9762 ± 0.0037 |
| 0.5 | TCGA | 0.9224 ± 0.0090 | 0.9739 ± 0.0040 | 0.9266 ± 0.0076 | **0.9764 ± 0.0037** |
| 0.6 | TCGA | 0.9216 ± 0.0095 | 0.9739 ± 0.0039 | 0.9264 ± 0.0080 | 0.9761 ± 0.0037 |
| 0.7 | TCGA | 0.9229 ± 0.0090 | 0.9744 ± 0.0038 | 0.9248 ± 0.0084 | 0.9761 ± 0.0037 |
| 0.8 | TCGA | 0.9224 ± 0.0086 | 0.9744 ± 0.0038 | 0.9253 ± 0.0083 | 0.9758 ± 0.0037 |
| 0.9 | TCGA | 0.9224 ± 0.0093 | 0.9747 ± 0.0038 | 0.9242 ± 0.0084 | 0.9753 ± 0.0038 |
| No mask | TCGA | 0.9224 ± 0.0089 | 0.9747 ± 0.0038 | 0.9224 ± 0.0089 | 0.9747 ± 0.0038 |

Supplementary Table 45: **Impact of mask ratio and elimination strategies on patient-level accuracy and AUROC of CONCH-SNGP without retraining regarding the TCGA test data**. In the following supplementary material, unless otherwise specified, the spectral normalization factor $c$ of CONCH-SNGP is fixed at 2.0. Best-performing model for each case is highlighted in bold, and results are presented with mean ± 95% CI from 20 experimental replicates.

| Mask Ratio | Evaluation dataset | **Random** | | **Ambiguity** | |
|---|---|---|---|---|---|
| | | Accuracy | AUROC | Accuracy | AUROC |
| 0.001 | CPTAC | 0.8679 ± 0.0148 | 0.9571 ± 0.0064 | 0.9512 ± 0.0017 | 0.9762 ± 0.0015 |
| 0.01 | CPTAC | 0.9327 ± 0.0058 | 0.9793 ± 0.0027 | 0.9500 ± 0.0023 | 0.9803 ± 0.0016 |
| 0.1 | CPTAC | 0.9446 ± 0.0052 | 0.9837 ± 0.0006 | 0.9518 ± 0.0013 | 0.9822 ± 0.0007 |
| 0.2 | CPTAC | 0.9440 ± 0.0044 | 0.9838 ± 0.0011 | **0.9524 ± 0.0000** | 0.9821 ± 0.0013 |
| 0.3 | CPTAC | 0.9494 ± 0.0036 | 0.9839 ± 0.0012 | **0.9524 ± 0.0000** | 0.9821 ± 0.0014 |
| 0.4 | CPTAC | 0.9500 ± 0.0029 | 0.9838 ± 0.0013 | **0.9524 ± 0.0000** | 0.9821 ± 0.0014 |
| 0.5 | CPTAC | 0.9488 ± 0.0032 | 0.9838 ± 0.0013 | **0.9524 ± 0.0000** | 0.9823 ± 0.0015 |
| 0.6 | CPTAC | 0.9476 ± 0.0028 | 0.9837 ± 0.0015 | **0.9524 ± 0.0000** | 0.9826 ± 0.0015 |
| 0.7 | CPTAC | 0.9488 ± 0.0026 | 0.9839 ± 0.0014 | **0.9524 ± 0.0000** | 0.9830 ± 0.0015 |
| 0.8 | CPTAC | 0.9482 ± 0.0027 | 0.9836 ± 0.0014 | **0.9524 ± 0.0000** | 0.9832 ± 0.0015 |
| 0.9 | CPTAC | 0.9476 ± 0.0028 | 0.9836 ± 0.0013 | 0.9506 ± 0.0021 | 0.9834 ± 0.0015 |
| No mask | CPTAC | 0.9476 ± 0.0028 | 0.9836 ± 0.0013 | 0.9476 ± 0.0028 | **0.9836 ± 0.0013** |

Supplementary Table 46: **Impact of mask ratio and elimination strategies on patient-level accuracy and AUROC of CONCH-SNGP without retraining regarding the CPTAC test data**. The same setting of UNI-SNGP described earlier was applied on CPTAC. Best-performing model for each case is highlighted in bold, and results are presented with mean ± 95% CI from 20 experimental replicates.

| Evaluation dataset | Mask Ratio | Average set size | |
|---|---|---|---|
| | | $\alpha = 0.05$ | $\alpha = 0.01$ |
| TCGA | 0.001 | 1.1279 ± 0.0029 | 1.5732 ± 0.0040 |
| TCGA | 0.01 | 1.0907 ± 0.0023 | 1.5574 ± 0.0040 |
| TCGA | 0.1 | 1.0727 ± 0.0020 | 1.5429 ± 0.0040 |
| TCGA | 0.2 | 1.0718 ± 0.0020 | 1.5378 ± 0.0041 |
| TCGA | 0.3 | 1.0693 ± 0.0019 | 1.5376 ± 0.0042 |
| TCGA | 0.4 | 1.0692 ± 0.0019 | 1.5383 ± 0.0042 |
| TCGA | 0.5 | **1.0690 ± 0.0018** | 1.5389 ± 0.0042 |
| TCGA | 0.6 | 1.0691 ± 0.0018 | 1.5398 ± 0.0042 |
| TCGA | 0.7 | 1.0695 ± 0.0018 | 1.5381 ± 0.0042 |
| TCGA | 0.8 | 1.0696 ± 0.0017 | 1.5366 ± 0.0042 |
| TCGA | 0.9 | 1.0709 ± 0.0017 | **1.5355 ± 0.0042** |
| TCGA | No mask | 1.0742 ± 0.0017 | 1.5360 ± 0.0042 |

Supplementary Table 47: **Impact of mask ratio on the average set size of UNI-SNGP-EAT without retraining (implemented with the ambiguity-based EAT) regarding the TCGA test data**. As the coverage for UNI-SNGP-EAT and CONCH-SNGP-EAT is guaranteed, we have omitted reporting their empirical coverage values. Best-performing model for each case is highlighted in bold, and results are presented with mean ± 95% CI from 20 experimental replicates with each using 500 randomly selected calibration/test permutations.

| Evaluation dataset | Mask Ratio | Average set size | |
|---|---|---|---|
| | | $\alpha = 0.05$ | $\alpha = 0.01$ |
| CPTAC | 0.001 | 1.1726 ± 0.0021 | 1.4979 ± 0.0042 |
| CPTAC | 0.01 | 1.1233 ± 0.0027 | 1.5283 ± 0.0045 |
| CPTAC | 0.1 | 1.0686 ± 0.0018 | 1.4896 ± 0.0043 |
| CPTAC | 0.2 | 1.0628 ± 0.0018 | 1.4843 ± 0.0043 |
| CPTAC | 0.3 | 1.0552 ± 0.0018 | 1.4820 ± 0.0043 |
| CPTAC | 0.4 | 1.0525 ± 0.0018 | 1.4812 ± 0.0043 |
| CPTAC | 0.5 | 1.0516 ± 0.0018 | 1.4781 ± 0.0043 |
| CPTAC | 0.6 | **1.0509 ± 0.0018** | **1.4768 ± 0.0042** |
| CPTAC | 0.7 | 1.0517 ± 0.0018 | 1.4810 ± 0.0042 |
| CPTAC | 0.8 | 1.0522 ± 0.0018 | 1.4820 ± 0.0042 |
| CPTAC | 0.9 | 1.0534 ± 0.0018 | 1.4829 ± 0.0042 |
| CPTAC | No mask | 1.0574 ± 0.0019 | 1.4870 ± 0.0041 |

Supplementary Table 48: **Impact of mask ratio on the average set size of UNI-SNGP-EAT without retraining (implemented with the ambiguity-based EAT) regarding the CPTAC test data**. Note that the CPTAC test set only contains 86 instances, making it impossible to construct both a calibration and test set for $\alpha = 0.01$. To address this issue, we bootstrap 100 samples and introduce a small amount of noise to break ties when creating the calibration set. Best-performing model for each case is highlighted in bold, and results are presented with mean ± 95% CI from 20 experimental replicates with each using 500 randomly selected calibration/test permutations.

| Evaluation dataset | Mask Ratio | Average set size | |
|---|---|---|---|
| | | $\alpha = 0.05$ | $\alpha = 0.01$ |
| TCGA | 0.001 | 1.1216 ± 0.0026 | 1.5752 ± 0.0044 |
| TCGA | 0.01 | 1.0986 ± 0.0022 | 1.5443 ± 0.0045 |
| TCGA | 0.1 | 1.0923 ± 0.0019 | 1.5351 ± 0.0048 |
| TCGA | 0.2 | **1.0913 ± 0.0018** | 1.5251 ± 0.0048 |
| TCGA | 0.3 | **1.0913 ± 0.0017** | 1.5148 ± 0.0047 |
| TCGA | 0.4 | 1.0916 ± 0.0017 | 1.5085 ± 0.0047 |
| TCGA | 0.5 | 1.0932 ± 0.0017 | **1.5051 ± 0.0047** |
| TCGA | 0.6 | 1.0953 ± 0.0017 | 1.5058 ± 0.0047 |
| TCGA | 0.7 | 1.0970 ± 0.0017 | 1.5062 ± 0.0048 |
| TCGA | 0.8 | 1.0992 ± 0.0017 | 1.5072 ± 0.0048 |
| TCGA | 0.9 | 1.1026 ± 0.0017 | 1.5074 ± 0.0048 |
| TCGA | No mask | 1.1068 ± 0.0016 | 1.5088 ± 0.0048 |

Supplementary Table 49: **Impact of mask ratio on the average set size of CONCH-SNGP-EAT without retraining (implemented with the ambiguity-based EAT) regarding the TCGA test data**. Best-performing model for each case is highlighted in bold, and results are presented with mean ± 95% CI from 20 experimental replicates with each using 500 randomly selected calibration/test permutations.

| Evaluation dataset | Mask Ratio | Average set size | |
|---|---|---|---|
| | | $\alpha = 0.05$ | $\alpha = 0.01$ |
| CPTAC | 0.001 | 1.0488 ± 0.0017 | 1.7683 ± 0.0057 |
| CPTAC | 0.01 | 1.0337 ± 0.0014 | 1.6600 ± 0.0051 |
| CPTAC | 0.1 | 1.0296 ± 0.0011 | 1.5894 ± 0.0049 |
| CPTAC | 0.2 | 1.0257 ± 0.0010 | 1.5424 ± 0.0046 |
| CPTAC | 0.3 | 1.0226 ± 0.0009 | 1.5210 ± 0.0046 |
| CPTAC | 0.4 | 1.0203 ± 0.0009 | 1.5152 ± 0.0046 |
| CPTAC | 0.5 | 1.0188 ± 0.0008 | 1.5143 ± 0.0046 |
| CPTAC | 0.6 | 1.0170 ± 0.0008 | 1.5112 ± 0.0046 |
| CPTAC | 0.7 | 1.0158 ± 0.0008 | 1.5112 ± 0.0046 |
| CPTAC | 0.8 | **1.0155 ± 0.0008** | 1.5113 ± 0.0045 |
| CPTAC | 0.9 | 1.0160 ± 0.0007 | **1.5098 ± 0.0045** |
| CPTAC | No mask | 1.0174 ± 0.0008 | 1.5156 ± 0.0045 |

Supplementary Table 50: **Impact of mask ratio on the average set size of CONCH-SNGP-EAT without retraining (implemented with the ambiguity-based EAT) regarding the CPTACT test data**. Best-performing model for each case is highlighted in bold, and results are presented with mean ± 95% CI from 20 experimental replicates with each using 500 randomly selected calibration/test permutations.

| Model | In-D | Mask Ratio | AUROC | | |
|---|---|---|---|---|---|
| | | | UCS | UVM | BLCA |
| CONCH-SNGP | TCGA | 0.001 | 0.9853 ± 0.0031 | 0.9931 ± 0.0016 | 0.9716 ± 0.0045 |
| CONCH-SNGP | TCGA | 0.01 | 0.9864 ± 0.0049 | 0.9949 ± 0.0018 | 0.9756 ± 0.0049 |
| CONCH-SNGP | TCGA | 0.1 | 0.9870 ± 0.0039 | 0.9957 ± 0.0016 | 0.9774 ± 0.0041 |
| CONCH-SNGP | TCGA | 0.2 | 0.9876 ± 0.0032 | 0.9960 ± 0.0015 | 0.9778 ± 0.0043 |
| CONCH-SNGP | TCGA | 0.3 | 0.9882 ± 0.0030 | 0.9964 ± 0.0015 | 0.9782 ± 0.0042 |
| CONCH-SNGP | TCGA | 0.4 | 0.9888 ± 0.0027 | 0.9966 ± 0.0015 | 0.9784 ± 0.0041 |
| CONCH-SNGP | TCGA | 0.5 | 0.9893 ± 0.0027 | 0.9968 ± 0.0013 | 0.9786 ± 0.0039 |
| CONCH-SNGP | TCGA | 0.6 | 0.9898 ± 0.0025 | 0.9969 ± 0.0012 | 0.9788 ± 0.0039 |
| CONCH-SNGP | TCGA | 0.7 | 0.9899 ± 0.0025 | 0.9970 ± 0.0012 | 0.9787 ± 0.0039 |
| CONCH-SNGP | TCGA | 0.8 | 0.9900 ± 0.0025 | 0.9970 ± 0.0014 | 0.9786 ± 0.0040 |
| CONCH-SNGP | TCGA | 0.9 | 0.9902 ± 0.0025 | 0.9970 ± 0.0014 | 0.9785 ± 0.0039 |
| CONCH-SNGP | TCGA | No mask | 0.9903 ± 0.0025 | 0.9970 ± 0.0013 | 0.9786 ± 0.0040 |

| Model | In-D | Mask Ratio | AUROC | |
|---|---|---|---|---|
| | | | ACC | Average |
| CONCH-SNGP | TCGA | 0.001 | 0.9920 ± 0.0018 | 0.9782 ± 0.0034 |
| CONCH-SNGP | TCGA | 0.01 | 0.9929 ± 0.0022 | 0.9812 ± 0.0038 |
| CONCH-SNGP | TCGA | 0.1 | 0.9930 ± 0.0020 | 0.9826 ± 0.0032 |
| CONCH-SNGP | TCGA | 0.2 | 0.9934 ± 0.0020 | 0.9830 ± 0.0033 |
| CONCH-SNGP | TCGA | 0.3 | 0.9937 ± 0.0020 | 0.9834 ± 0.0032 |
| CONCH-SNGP | TCGA | 0.4 | 0.9941 ± 0.0020 | 0.9836 ± 0.0031 |
| CONCH-SNGP | TCGA | 0.5 | 0.9944 ± 0.0019 | 0.9839 ± 0.0030 |
| CONCH-SNGP | TCGA | 0.6 | 0.9945 ± 0.0019 | **0.9841 ± 0.0030** |
| CONCH-SNGP | TCGA | 0.7 | 0.9945 ± 0.0019 | 0.9840 ± 0.0030 |
| CONCH-SNGP | TCGA | 0.8 | 0.9944 ± 0.0021 | 0.9840 ± 0.0031 |
| CONCH-SNGP | TCGA | 0.9 | 0.9944 ± 0.0021 | 0.9839 ± 0.0031 |
| CONCH-SNGP | TCGA | No mask | 0.9946 ± 0.0021 | 0.9840 ± 0.0031 |

Supplementary Table 51: **Impact of mask ratio on patient-level fine-grained OOD detection performance of CONCH with In-D TCGA**. Performance results for various mask ratio of CONCH-SNGP-EAT are reported. Best-performing model for each case is highlighted in bold, and results are presented with mean ± 95% CI from 20 experimental replicates.

| Model | In-D | Mask Ratio | AUROC | | |
|---|---|---|---|---|---|
| | | | UCS | UVM | BLCA |
| CONCH-SNGP | CPTAC | 0.001 | 0.9423 ± 0.0080 | 0.9843 ± 0.0032 | 0.8719 ± 0.0137 |
| CONCH-SNGP | CPTAC | 0.01 | 0.9247 ± 0.0124 | 0.9938 ± 0.0013 | 0.8831 ± 0.0139 |
| CONCH-SNGP | CPTAC | 0.1 | 0.9397 ± 0.0090 | 0.9950 ± 0.0010 | 0.9053 ± 0.0105 |
| CONCH-SNGP | CPTAC | 0.2 | 0.9505 ± 0.0067 | 0.9961 ± 0.0007 | 0.9151 ± 0.0083 |
| CONCH-SNGP | CPTAC | 0.3 | 0.9564 ± 0.0056 | 0.9965 ± 0.0007 | 0.9207 ± 0.0070 |
| CONCH-SNGP | CPTAC | 0.4 | 0.9601 ± 0.0046 | 0.9967 ± 0.0006 | 0.9241 ± 0.0064 |
| CONCH-SNGP | CPTAC | 0.5 | 0.9629 ± 0.0042 | 0.9970 ± 0.0006 | 0.9271 ± 0.0059 |
| CONCH-SNGP | CPTAC | 0.6 | 0.9652 ± 0.0038 | 0.9972 ± 0.0006 | 0.9292 ± 0.0055 |
| CONCH-SNGP | CPTAC | 0.7 | 0.9672 ± 0.0037 | 0.9974 ± 0.0006 | 0.9312 ± 0.0051 |
| CONCH-SNGP | CPTAC | 0.8 | 0.9688 ± 0.0035 | 0.9976 ± 0.0005 | 0.9331 ± 0.0050 |
| CONCH-SNGP | CPTAC | 0.9 | 0.9698 ± 0.0036 | 0.9979 ± 0.0004 | 0.9347 ± 0.0049 |
| CONCH-SNGP | CPTAC | No mask | 0.9713 ± 0.0036 | 0.9980 ± 0.0004 | 0.9363 ± 0.0049 |

| Model | In-D | Mask Ratio | AUROC | |
|---|---|---|---|---|
| | | | ACC | Average |
| CONCH-SNGP | CPTAC | 0.001 | 0.9682 ± 0.0053 | 0.9048 ± 0.0104 |
| CONCH-SNGP | CPTAC | 0.01 | 0.9618 ± 0.0078 | 0.9106 ± 0.0111 |
| CONCH-SNGP | CPTAC | 0.1 | 0.9717 ± 0.0042 | 0.9280 ± 0.0082 |
| CONCH-SNGP | CPTAC | 0.2 | 0.9793 ± 0.0029 | 0.9367 ± 0.0063 |
| CONCH-SNGP | CPTAC | 0.3 | 0.9833 ± 0.0022 | 0.9415 ± 0.0053 |
| CONCH-SNGP | CPTAC | 0.4 | 0.9855 ± 0.0018 | 0.9444 ± 0.0047 |
| CONCH-SNGP | CPTAC | 0.5 | 0.9874 ± 0.0015 | 0.9469 ± 0.0043 |
| CONCH-SNGP | CPTAC | 0.6 | 0.9887 ± 0.0013 | 0.9487 ± 0.0040 |
| CONCH-SNGP | CPTAC | 0.7 | 0.9897 ± 0.0012 | 0.9503 ± 0.0037 |
| CONCH-SNGP | CPTAC | 0.8 | 0.9904 ± 0.0012 | 0.9518 ± 0.0037 |
| CONCH-SNGP | CPTAC | 0.9 | 0.9912 ± 0.0011 | 0.9531 ± 0.0036 |
| CONCH-SNGP | CPTAC | No mask | 0.9921 ± 0.0010 | **0.9545 ± 0.0036** |

Supplementary Table 52: **Impact of mask ratio on patient-level fine-grained OOD detection performance of CONCH with In-D CPTAC**. Performance results for various mask ratio of CONCH-SNGP-EAT are reported. Best-performing model for each case is highlighted in bold, and results are presented with mean ± 95% CI from 20 experimental replicates.

| Model | Metric | Not reported | Others | White | Fairness gap |
|---|---|---|---|---|---|
| UNI | Accuracy | 0.9467 ± 0.0158 | 0.8722 ± 0.0264 | 0.9352 ± 0.0066 | 0.0891 ± 0.0264 |
| UNI-SNGP | Accuracy | 0.9560 ± 0.0163 | 0.9006 ± 0.0249 | 0.9379 ± 0.0080 | **0.0759 ± 0.0218** |
| UNI-SNGP-RE | Accuracy | 0.9558 ± 0.0146 | 0.8888 ± 0.0233 | 0.9368 ± 0.0086 | 0.0790 ± 0.0235 |
| UNI-SNGP-EAT | Accuracy | 0.9555 ± 0.0138 | 0.8985 ± 0.0257 | 0.9391 ± 0.0081 | 0.0774 ± 0.0234 |
| UNI | AUROC | 0.9710 ± 0.0103 | 0.9323 ± 0.0271 | 0.9813 ± 0.0037 | 0.0610 ± 0.0254 |
| UNI-SNGP | AUROC | 0.9590 ± 0.0162 | 0.9469 ± 0.0183 | 0.9800 ± 0.0042 | 0.0595 ± 0.0162 |
| UNI-SNGP-RE | AUROC | 0.9593 ± 0.0155 | 0.9476 ± 0.0188 | 0.9800 ± 0.0040 | **0.0589 ± 0.0165** |
| UNI-SNGP-EAT | AUROC | 0.9580 ± 0.0165 | 0.9512 ± 0.0181 | 0.9799 ± 0.0043 | 0.0590 ± 0.0155 |

| Model | Metric | Not reported | Others | White | Fairness gap |
|---|---|---|---|---|---|
| CONCH | Accuracy | 0.9220 ± 0.0269 | 0.8654 ± 0.0246 | 0.9297 ± 0.0099 | 0.0949 ± 0.0257 |
| CONCH-SNGP | Accuracy | 0.9249 ± 0.0235 | 0.8779 ± 0.0264 | 0.9274 ± 0.0112 | **0.0799 ± 0.0229** |
| CONCH-SNGP-RE | Accuracy | 0.9232 ± 0.0241 | 0.8729 ± 0.0288 | 0.9285 ± 0.0118 | 0.0903 ± 0.0250 |
| CONCH-SNGP-EAT | Accuracy | 0.9321 ± 0.0227 | 0.8782 ± 0.0271 | 0.9327 ± 0.0091 | 0.0813 ± 0.0251 |
| CONCH | AUROC | 0.9584 ± 0.0141 | 0.9470 ± 0.0192 | 0.9806 ± 0.0047 | 0.0552 ± 0.0180 |
| CONCH-SNGP | AUROC | 0.9608 ± 0.0122 | 0.9516 ± 0.0165 | 0.9826 ± 0.0044 | 0.0507 ± 0.0153 |
| CONCH-SNGP-RE | AUROC | 0.9605 ± 0.0138 | 0.9502 ± 0.0173 | 0.9818 ± 0.0048 | 0.0525 ± 0.0166 |
| CONCH-SNGP-EAT | AUROC | 0.9661 ± 0.0115 | 0.9502 ± 0.0183 | 0.9836 ± 0.0042 | **0.0496 ± 0.0176** |

Supplementary Table 53: **Patient-level race-wise fairness gap (Not reported (n=36), Others (n=20), White (n=133)) of UNI, CONCH and their variants with respect to the TCGA test data.** Note that race categories with fewer than 20 patients are aggregated as 'Others'. Best-performing model for each case is highlighted in bold, and results are presented with mean ± 95% CI from 20 experimental replicates.

| Model | Metric | Asian | Others | White | Fairness gap |
|---|---|---|---|---|---|
| UNI | Accuracy | 1.0000 ± 0.0000 | 0.9000 ± 0.0000 | 0.9211 ± 0.0052 | 0.1000 ± 0.0000 |
| UNI-SNGP | Accuracy | 1.0000 ± 0.0000 | 0.8950 ± 0.0102 | 0.9222 ± 0.0070 | 0.1061 ± 0.0102 |
| UNI-SNGP-RE | Accuracy | 0.9983 ± 0.0035 | 0.9000 ± 0.0000 | 0.9200 ± 0.0061 | **0.0988 ± 0.0037** |
| UNI-SNGP-EAT | Accuracy | 1.0000 ± 0.0000 | 0.9000 ± 0.0000 | 0.9233 ± 0.0061 | 0.1006 ± 0.0011 |
| UNI | AUROC | 1.0000 ± 0.0000 | 0.9792 ± 0.0097 | 0.9568 ± 0.0017 | **0.0434 ± 0.0016** |
| UNI-SNGP | AUROC | 1.0000 ± 0.0000 | 0.9708 ± 0.0139 | 0.9578 ± 0.0035 | 0.0504 ± 0.0071 |
| UNI-SNGP-RE | AUROC | 1.0000 ± 0.0000 | 0.9562 ± 0.0157 | 0.9591 ± 0.0026 | 0.0576 ± 0.0090 |
| UNI-SNGP-EAT | AUROC | 1.0000 ± 0.0000 | 0.9729 ± 0.0127 | 0.9582 ± 0.0040 | 0.0483 ± 0.0063 |

| Model | Metric | Asian | Others | White | Fairness gap |
|---|---|---|---|---|---|
| CONCH | Accuracy | 0.9966 ± 0.0048 | 0.9000 ± 0.0000 | 0.9267 ± 0.0048 | **0.0966 ± 0.0048** |
| CONCH-SNGP | Accuracy | 1.0000 ± 0.0000 | 0.9000 ± 0.0000 | 0.9267 ± 0.0048 | 0.1000 ± 0.0000 |
| CONCH-SNGP-RE | Accuracy | 1.0000 ± 0.0000 | 0.9000 ± 0.0000 | 0.9278 ± 0.0065 | 0.1006 ± 0.0011 |
| CONCH-SNGP-EAT | Accuracy | 1.0000 ± 0.0000 | 0.9000 ± 0.0000 | 0.9333 ± 0.0000 | 0.1000 ± 0.0000 |
| CONCH | AUROC | 1.0000 ± 0.0000 | 0.9156 ± 0.0176 | 0.9568 ± 0.0029 | 0.0844 ± 0.0176 |
| CONCH-SNGP | AUROC | 1.0000 ± 0.0000 | 0.9604 ± 0.0043 | 0.9642 ± 0.0010 | 0.0415 ± 0.0003 |
| CONCH-SNGP-RE | AUROC | 1.0000 ± 0.0000 | 0.9604 ± 0.0043 | 0.9654 ± 0.0017 | **0.0414 ± 0.0005** |
| CONCH-SNGP-EAT | AUROC | 1.0000 ± 0.0000 | 0.9583 ± 0.0000 | 0.9606 ± 0.0011 | 0.0420 ± 0.0007 |

Supplementary Table 54: **Patient-level race-wise fairness gap (Asian (n=29), Others (n=10), White (n=45)) of UNI, CONCH and their variants with respect to the CPTAC test data.** Best-performing model for each case is highlighted in bold, and results are presented with mean ± 95% CI from 20 experimental replicates.

| Model | Metric | Female | Male | Fairness gap |
|---|---|---|---|---|
| UNI | Accuracy | 0.9244 ± 0.0102 | 0.9346 ± 0.0067 | **0.0244 ± 0.0064** |
| UNI-SNGP | Accuracy | 0.9472 ± 0.0126 | 0.9324 ± 0.0082 | 0.0281 ± 0.0107 |
| UNI-SNGP-RE | Accuracy | 0.9475 ± 0.0130 | 0.9291 ± 0.0085 | 0.0326 ± 0.0095 |
| UNI-SNGP-EAT | Accuracy | 0.9473 ± 0.0125 | 0.9331 ± 0.0078 | 0.0294 ± 0.0093 |
| UNI | AUROC | 0.9783 ± 0.0060 | 0.9696 ± 0.0058 | 0.0165 ± 0.0062 |
| UNI-SNGP | AUROC | 0.9773 ± 0.0066 | 0.9693 ± 0.0041 | 0.0131 ± 0.0059 |
| UNI-SNGP-RE | AUROC | 0.9774 ± 0.0066 | 0.9694 ± 0.0039 | 0.0137 ± 0.0056 |
| UNI-SNGP-EAT | AUROC | 0.9777 ± 0.0066 | 0.9699 ± 0.0039 | **0.0125 ± 0.0058** |

| Model | Metric | Female | Male | Fairness gap |
|---|---|---|---|---|
| CONCH | Accuracy | 0.9207 ± 0.0141 | 0.9220 ± 0.0067 | 0.0228 ± 0.0070 |
| CONCH-SNGP | Accuracy | 0.9229 ± 0.0150 | 0.9214 ± 0.0090 | 0.0242 ± 0.0118 |
| CONCH-SNGP-RE | Accuracy | 0.9267 ± 0.0148 | 0.9193 ± 0.0102 | **0.0221 ± 0.0124** |
| CONCH-SNGP-EAT | Accuracy | 0.9266 ± 0.0133 | 0.9269 ± 0.0077 | 0.0262 ± 0.0080 |
| CONCH | AUROC | 0.9772 ± 0.0051 | 0.9683 ± 0.0060 | 0.0135 ± 0.0047 |
| CONCH-SNGP | AUROC | 0.9793 ± 0.0058 | 0.9699 ± 0.0052 | 0.0131 ± 0.0050 |
| CONCH-SNGP-RE | AUROC | 0.9779 ± 0.0065 | 0.9695 ± 0.0054 | **0.0130 ± 0.0052** |
| CONCH-SNGP-EAT | AUROC | 0.9811 ± 0.0059 | 0.9714 ± 0.0049 | 0.0138 ± 0.0046 |

Supplementary Table 55: **Patient-level sex-wise fairness gap among different groups (Female (n=68), Male (n=121)) of UNI, CONCH and their variants with respect to the TCGA test data.** Best-performing model for each case is highlighted in bold, and results are presented with mean ± 95% CI from 20 experimental replicates.

| Model | Metric | Female | Male | Fairness gap |
|---|---|---|---|---|
| UNI | Accuracy | 0.9259 ± 0.0000 | 0.9553 ± 0.0041 | **0.0293 ± 0.0041** |
| UNI-SNGP | Accuracy | 0.9167 ± 0.0075 | 0.9596 ± 0.0037 | 0.0430 ± 0.0080 |
| UNI-SNGP-RE | Accuracy | 0.9148 ± 0.0080 | 0.9588 ± 0.0039 | 0.0440 ± 0.0097 |
| UNI-SNGP-EAT | Accuracy | 0.9167 ± 0.0075 | 0.9614 ± 0.0033 | 0.0447 ± 0.0082 |
| UNI | AUROC | 0.9671 ± 0.0000 | 0.9760 ± 0.0008 | **0.0088 ± 0.0008** |
| UNI-SNGP | AUROC | 0.9622 ± 0.0027 | 0.9805 ± 0.0019 | 0.0184 ± 0.0036 |
| UNI-SNGP-RE | AUROC | 0.9625 ± 0.0024 | 0.9805 ± 0.0020 | 0.0180 ± 0.0035 |
| UNI-SNGP-EAT | AUROC | 0.9599 ± 0.0052 | 0.9808 ± 0.0021 | 0.0210 ± 0.0061 |

| Model | Metric | Female | Male | Fairness gap |
|---|---|---|---|---|
| CONCH | Accuracy | 0.9222 ± 0.0052 | 0.9596 ± 0.0037 | 0.0374 ± 0.0071 |
| CONCH-SNGP | Accuracy | 0.9259 ± 0.0000 | 0.9596 ± 0.0037 | 0.0337 ± 0.0037 |
| CONCH-SNGP-RE | Accuracy | 0.9278 ± 0.0038 | 0.9596 ± 0.0046 | **0.0319 ± 0.0056** |
| CONCH-SNGP-EAT | Accuracy | 0.9259 ± 0.0000 | 0.9649 ± 0.0000 | 0.0390 ± 0.0000 |
| CONCH | AUROC | 0.9862 ± 0.0029 | 0.9619 ± 0.0066 | 0.0242 ± 0.0040 |
| CONCH-SNGP | AUROC | 0.9934 ± 0.0000 | 0.9786 ± 0.0022 | 0.0148 ± 0.0022 |
| CONCH-SNGP-RE | AUROC | 0.9934 ± 0.0000 | 0.9788 ± 0.0022 | 0.0146 ± 0.0022 |
| CONCH-SNGP-EAT | AUROC | 0.9872 ± 0.0012 | 0.9792 ± 0.0022 | **0.0080 ± 0.0017** |

Supplementary Table 56: **Patient-level sex-wise fairness gap (Female (n=27), Male (n=57)) of UNI, CONCH and their variants with respect to the CPTAC test data.** Best-performing model for each case is highlighted in bold, and results are presented with mean ± 95% CI from 20 experimental replicates.

| **Model** | $\alpha$ | **Average set size across racial groups** | | | |
|---|---|---|---|---|---|
| | | Not reported | Others | White | Fairness gap |
| UNI | 0.05 | 1.0557 ± 0.0018 | 1.0984 ± 0.0028 | 1.0907 ± 0.0017 | 0.1128 ± 0.0017 |
| UNI-SNGP | 0.05 | 1.0446 ± 0.0014 | 1.0891 ± 0.0028 | 1.0807 ± 0.0018 | 0.1044 ± 0.0018 |
| UNI-SNGP-RE | 0.05 | 1.0441 ± 0.0014 | 1.0912 ± 0.0027 | 1.0853 ± 0.0018 | 0.1070 ± 0.0018 |
| UNI-SNGP-EAT | 0.05 | 1.0355 ± 0.0014 | 1.0811 ± 0.0028 | 1.0772 ± 0.0020 | **0.1006 ± 0.0019** |
| UNI | 0.01 | 1.5616 ± 0.0060 | 1.6155 ± 0.0051 | 1.5731 ± 0.0048 | **0.1809 ± 0.0022** |
| UNI-SNGP | 0.01 | 1.4562 ± 0.0054 | 1.5601 ± 0.0046 | 1.5529 ± 0.0041 | 0.2304 ± 0.0029 |
| UNI-SNGP-RE | 0.01 | 1.4498 ± 0.0053 | 1.5518 ± 0.0046 | 1.5580 ± 0.0041 | 0.2348 ± 0.0029 |
| UNI-SNGP-EAT | 0.01 | 1.4480 ± 0.0055 | 1.5603 ± 0.0046 | 1.5588 ± 0.0041 | 0.2355 ± 0.0029 |

| **Model** | $\alpha$ | **Average set size across racial groups** | | | |
|---|---|---|---|---|---|
| | | Not reported | Others | White | Fairness gap |
| CONCH | 0.05 | 1.0821 ± 0.0019 | 1.1362 ± 0.0028 | 1.1218 ± 0.0017 | 0.1308 ± 0.0018 |
| CONCH-SNGP | 0.05 | 1.0776 ± 0.0019 | 1.1266 ± 0.0026 | 1.1116 ± 0.0017 | 0.1225 ± 0.0017 |
| CONCH-SNGP-RE | 0.05 | 1.0833 ± 0.0019 | 1.1368 ± 0.0027 | 1.1178 ± 0.0018 | 0.1262 ± 0.0017 |
| CONCH-SNGP-EAT | 0.05 | 1.0536 ± 0.0018 | 1.1121 ± 0.0026 | 1.0989 ± 0.0018 | **0.1158 ± 0.0017** |
| CONCH | 0.01 | 1.5017 ± 0.0058 | 1.5811 ± 0.0054 | 1.5551 ± 0.0051 | **0.1740 ± 0.0021** |
| CONCH-SNGP | 0.01 | 1.4547 ± 0.0052 | 1.5358 ± 0.0051 | 1.5193 ± 0.0049 | 0.1780 ± 0.0021 |
| CONCH-SNGP-RE | 0.01 | 1.4580 ± 0.0052 | 1.5380 ± 0.0051 | 1.5223 ± 0.0049 | 0.1782 ± 0.0021 |
| CONCH-SNGP-EAT | 0.01 | 1.4380 ± 0.0053 | 1.5466 ± 0.0051 | 1.5218 ± 0.0047 | 0.1935 ± 0.0022 |

Supplementary Table 57: **Patient-level race-wise fairness gap on average set size (Not reported (n=36), Others (n=20), White (n=133)) of UNI, CONCH and their variants with respect to the internal TCGA test data**. Best-performing model for fairness gap is highlighted in bold, and results are presented with mean ± 95% confidence interval (CI) from 20 experimental replicates.

| **Model** | $\alpha$ | **Average set size across racial groups** | | | |
|---|---|---|---|---|---|
| | | Asian | Others | White | Fairness gap |
| UNI | 0.05 | 1.0319 ± 0.0009 | 1.1053 ± 0.0031 | 1.0633 ± 0.0015 | 0.1109 ± 0.0023 |
| UNI-SNGP | 0.05 | 1.0389 ± 0.0022 | 1.0809 ± 0.0030 | 1.0632 ± 0.0018 | 0.0991 ± 0.0023 |
| UNI-SNGP-RE | 0.05 | 1.0404 ± 0.0022 | 1.0940 ± 0.0033 | 1.0659 ± 0.0018 | 0.1072 ± 0.0025 |
| UNI-SNGP-EAT | 0.05 | 1.0317 ± 0.0022 | 1.0738 ± 0.0028 | 1.0600 ± 0.0018 | **0.0962 ± 0.0022** |
| UNI | 0.01 | 1.4113 ± 0.0029 | 1.5941 ± 0.0037 | 1.4413 ± 0.0031 | **0.2298 ± 0.0023** |
| UNI-SNGP | 0.01 | 1.3752 ± 0.0058 | 1.7105 ± 0.0041 | 1.5027 ± 0.0040 | 0.3984 ± 0.0032 |
| UNI-SNGP-RE | 0.01 | 1.3607 ± 0.0058 | 1.6896 ± 0.0044 | 1.4849 ± 0.0040 | 0.3934 ± 0.0032 |
| UNI-SNGP-EAT | 0.01 | 1.3294 ± 0.0062 | 1.7195 ± 0.0042 | 1.5160 ± 0.0042 | 0.4443 ± 0.0037 |

| **Model** | $\alpha$ | **Average set size across racial groups** | | | |
|---|---|---|---|---|---|
| | | Asian | Others | White | Fairness gap |
| CONCH | 0.05 | 1.0295 ± 0.0014 | 1.0077 ± 0.0009 | 1.0333 ± 0.0014 | 0.0592 ± 0.0011 |
| CONCH-SNGP | 0.05 | 1.0088 ± 0.0007 | 1.0005 ± 0.0002 | 1.0258 ± 0.0011 | **0.0490 ± 0.0008** |
| CONCH-SNGP-RE | 0.05 | 1.0083 ± 0.0007 | 1.0014 ± 0.0003 | 1.0256 ± 0.0011 | 0.0497 ± 0.0008 |
| CONCH-SNGP-EAT | 0.05 | 1.0030 ± 0.0004 | 1.0276 ± 0.0013 | 1.0291 ± 0.0013 | 0.0571 ± 0.0012 |
| CONCH | 0.01 | 1.6627 ± 0.0047 | 1.6311 ± 0.0049 | 1.6675 ± 0.0046 | 0.1743 ± 0.0019 |
| CONCH-SNGP | 0.01 | 1.5386 ± 0.0052 | 1.5125 ± 0.0051 | 1.5035 ± 0.0043 | **0.1632 ± 0.0019** |
| CONCH-SNGP-RE | 0.01 | 1.5378 ± 0.0052 | 1.5101 ± 0.0051 | 1.4996 ± 0.0043 | 0.1638 ± 0.0019 |
| CONCH-SNGP-EAT | 0.01 | 1.5184 ± 0.0056 | 1.4934 ± 0.0047 | 1.5180 ± 0.0043 | 0.1708 ± 0.0019 |

Supplementary Table 58: **Patient-level race-wise fairness gap on average set size (Asian (n=29), Others (n=10), White (n=45)) of UNI, CONCH and their variants with respect to the CPTAC test data**. Best-performing model for fairness is highlighted in bold, and results are presented with mean ± 95% confidence interval (CI) from 20 experimental replicates.

| Model | $\alpha$ | Average set size across sexual groups | | |
|---|---|---|---|---|
| | | Female | Male | Fairness gap |
| UNI | 0.05 | 1.0951 ± 0.0018 | 1.0781 ± 0.0018 | 0.0475 ± 0.0009 |
| UNI-SNGP | 0.05 | 1.0886 ± 0.0022 | 1.0660 ± 0.0015 | 0.0462 ± 0.0010 |
| UNI-SNGP-RE | 0.05 | 1.0911 ± 0.0021 | 1.0699 ± 0.0015 | 0.0470 ± 0.0010 |
| UNI-SNGP-EAT | 0.05 | 1.0846 ± 0.0024 | 1.0606 ± 0.0016 | **0.0457 ± 0.0011** |
| UNI | 0.01 | 1.5794 ± 0.0047 | 1.5740 ± 0.0054 | 0.1151 ± 0.0016 |
| UNI-SNGP | 0.01 | 1.6175 ± 0.0043 | 1.4894 ± 0.0045 | **0.1658 ± 0.0023** |
| UNI-SNGP-RE | 0.01 | 1.6200 ± 0.0043 | 1.4907 ± 0.0044 | 0.1691 ± 0.0023 |
| UNI-SNGP-EAT | 0.01 | 1.6288 ± 0.0042 | 1.4868 ± 0.0045 | 0.1786 ± 0.0022 |

| Model | $\alpha$ | Average set size across sexual groups | | |
|---|---|---|---|---|
| | | Female | Male | Fairness gap |
| CONCH | 0.05 | 1.1296 ± 0.0022 | 1.1077 ± 0.0015 | 0.0591 ± 0.0011 |
| CONCH-SNGP | 0.05 | 1.1189 ± 0.0023 | 1.0996 ± 0.0015 | 0.0576 ± 0.0011 |
| CONCH-SNGP-RE | 0.05 | 1.1242 ± 0.0023 | 1.1066 ± 0.0015 | 0.0583 ± 0.0011 |
| CONCH-SNGP-EAT | 0.05 | 1.1039 ± 0.0024 | 1.0847 ± 0.0015 | **0.0545 ± 0.0012** |
| CONCH | 0.01 | 1.5856 ± 0.0053 | 1.5265 ± 0.0053 | 0.1024 ± 0.0016 |
| CONCH-SNGP | 0.01 | 1.5497 ± 0.0051 | 1.4856 ± 0.0048 | 0.1024 ± 0.0016 |
| CONCH-SNGP-RE | 0.01 | 1.5549 ± 0.0051 | 1.4873 ± 0.0049 | **0.1022 ± 0.0016** |
| CONCH-SNGP-EAT | 0.01 | 1.5668 ± 0.0050 | 1.4757 ± 0.0048 | 0.1150 ± 0.0017 |

Supplementary Table 59: **Patient-level sex-wise fairness gap on average set size (Female (n=68), Male (n=121)) of UNI, CONCH and their variants with respect to the TCGA test data**. Best-performing model for fairness gap is highlighted in bold, and results are presented with mean ± 95% CI from 20 experimental replicates.

| Model | $\alpha$ | Average set size across sexual groups | | |
|---|---|---|---|---|
| | | Female | Male | Fairness gap |
| UNI | 0.05 | 1.0660 ± 0.0016 | 1.0541 ± 0.0014 | **0.0417 ± 0.0007** |
| UNI-SNGP | 0.05 | 1.0783 ± 0.0026 | 1.0469 ± 0.0016 | 0.0507 ± 0.0011 |
| UNI-SNGP-RE | 0.05 | 1.0788 ± 0.0025 | 1.0520 ± 0.0017 | 0.0482 ± 0.0011 |
| UNI-SNGP-EAT | 0.05 | 1.0726 ± 0.0025 | 1.0425 ± 0.0015 | 0.0561 ± 0.0011 |
| UNI | 0.01 | 1.3984 ± 0.0036 | 1.4755 ± 0.0028 | **0.1252 ± 0.0017** |
| UNI-SNGP | 0.01 | 1.4711 ± 0.0051 | 1.4948 ± 0.0040 | 0.1375 ± 0.0020 |
| UNI-SNGP-RE | 0.01 | 1.4437 ± 0.0049 | 1.4829 ± 0.0041 | 0.1304 ± 0.0019 |
| UNI-SNGP-EAT | 0.01 | 1.4914 ± 0.0050 | 1.4758 ± 0.0042 | 0.1296 ± 0.0018 |

| Model | $\alpha$ | Average set size across sexual groups | | |
|---|---|---|---|---|
| | | Female | Male | Fairness gap |
| CONCH | 0.05 | 1.0255 ± 0.0014 | 1.0310 ± 0.0013 | 0.0351 ± 0.0007 |
| CONCH-SNGP | 0.05 | 1.0084 ± 0.0007 | 1.0218 ± 0.0009 | 0.0273 ± 0.0005 |
| CONCH-SNGP-RE | 0.05 | 1.0079 ± 0.0008 | 1.0218 ± 0.0009 | 0.0286 ± 0.0005 |
| CONCH-SNGP-EAT | 0.05 | 1.0137 ± 0.0008 | 1.0235 ± 0.0010 | **0.0259 ± 0.0006** |
| CONCH | 0.01 | 1.6261 ± 0.0046 | 1.6796 ± 0.0047 | 0.1264 ± 0.0015 |
| CONCH-SNGP | 0.01 | 1.5042 ± 0.0053 | 1.5213 ± 0.0043 | 0.1070 ± 0.0015 |
| CONCH-SNGP-RE | 0.01 | 1.4945 ± 0.0052 | 1.5221 ± 0.0044 | **0.1049 ± 0.0015** |
| CONCH-SNGP-EAT | 0.01 | 1.5172 ± 0.0055 | 1.5142 ± 0.0043 | 0.1184 ± 0.0017 |

Supplementary Table 60: **Patient-level sex-wise performance fairness gap on average set size (Female (n=27), Male (n=57)) of UNI, CONCH and their variants with respect to the CPTAC test data**. Best-performing model for fairness gap is highlighted in bold, and results are presented with mean ± 95% CI from 20 experimental replicates with 500 calibration/test permutations.

| Dataset | | Link |
|---|---|---|
| TCGA | | https://portal.gdc.cancer.gov/projects/TCGA-LUAD<br>https://portal.gdc.cancer.gov/projects/TCGA-LUSC |
| CPTAC | | https://portal.gdc.cancer.gov/projects/CPTAC-3 |
| TCGA-OOD | UCS | https://portal.gdc.cancer.gov/projects/TCGA-UCS |
| | UVM | https://portal.gdc.cancer.gov/projects/TCGA-UVM |
| | BLCA | https://portal.gdc.cancer.gov/projects/TCGA-BLCA |
| | ACC | https://portal.gdc.cancer.gov/projects/TCGA-ACC |

Supplementary Table 61: **Summary of publicly available datasets.**
